# Computational analysis of peripheral blood smears detects disease-associated cytomorphologies

**DOI:** 10.1101/2022.04.19.22273757

**Authors:** José Guilherme de Almeida, Emma Gudgin, Martin Besser, William G. Dunn, Jonathan Cooper, Torsten Haferlach, George S. Vassiliou, Moritz Gerstung

**Author notes:** Correspondence to G.S.V, or M.G.

## Abstract

Many hematological diseases are characterized by altered abundance and morphology of blood cells and their progenitors. Myelodysplastic syndromes (MDS), for example, are a type of blood cancer manifesting via a range of cytopenias and dysplastic changes of blood and bone marrow cells. While experts analyze cytomorphology to diagnose MDS, similar alterations can be observed in other conditions such as haematinic deficiency anemias, and definitive diagnosis requires complementary information such as blood counts, karyotype and molecular testing. However, recent works demonstrated that computational analysis of bone marrow slides predicts not only MDS or AML but also the presence of specific mutations. Here, we present and make available Haemorasis, a computational method that detects and characterizes white and red blood cells (WBC and RBC, respectively) in peripheral blood slides, and apply it to over 300 individuals with different conditions (*SF3B1*-mutant and *SF3B1*-wildtype MDS, megaloblastic anemia and iron deficiency anemia), where Haemorasis detects over half a million WBC and millions of RBC. We then show how these large sets of cell images can be used in diagnosis and prognosis, whilst identifying novel associations between computational morphotypes and disease. We find that hypolobulated neutrophils and large RBC are characteristic of *SF3B1*-mutant MDS, and, while prevalent in both iron deficiency and megaloblastic anemia, hyperlobulated neutrophils are larger in the latter. Finally, we externally validate these methods, showing they generalize to other centers and scanners.

## 1 Introduction

The diagnosis of hematological malignancies relies on expert cytomorphological examination of blood, bone marrow and/or other tissue biopsies, together with molecular analyses that aid subclassification and prognosis ^1^. For example, anemias, characterized by reduced hemoglobin concentration (Hb) and red blood cell (RBC) numbers, can be both a disease and a feature of other conditions such as myelodysplastic syndromes (MDS), a heterogeneous group of myeloid neoplasms that can progress to acute myeloid leukemia (AML) ^2–4^. For this reason, the diagnosis of MDS requires the detection of cytopenias, changes to white blood cell (WBC) and RBC maturation through the analysis of blood cell morphology in bone marrow (BM) and peripheral blood slides (PBS), cyto- and histochemistry, karyotyping and immunophenotyping ^4–8^.

An accurate diagnosis of MDS and other hematological malignancies is essential to guide treatment — while megaloblastic anemia, which can be confounded with MDS ^9–11^, is generally treated with dietary changes or supplements ^12^, the treatment of MDS generally involves chemotherapeutic agents or blood/platelet transfusions ^13^ and depends on risk assessments which consider blood counts, BM cytomorphology and cytogenetics ^14^. Furthermore, MDS prognosis can also benefit from molecular characterization, used to define clinically-relevant MDS subtypes such as *SF3B1*-mutant MDS, associated with improved survival times ^15,16^. It should be noted that MDS cases with splicing factor mutations such as *SF3B1*-mutant MDS account for over 50% of MDS cases ^17,18^, constituting an important MDS subtype.

While abnormalities such as an increased prevalence of hypolobulated granulocytes, abnormal granularity in neutrophils or abnormal RBC have been observed in MDS ^8,19–21^, peripheral blood cell morphology is generally insufficient for MDS diagnosis. This is also compounded by challenges associated with the assessment of subtle cytomorphological alterations and heterogeneity across a given slide leading to inter-observer variation. While diagnoses stemming from the analysis of a whole PBS (requiring the analysis of hundreds of cells) typically show high concordance, the classification and characterization of individual WBC is more challenging ^22,23^. Additionally, the evidence on whether trained experts can distinguish specific cell types, particularly monocyte subtypes, is conflicting ^24,25^, and a study looking specifically at cell type classification concordance among 28 morphologists identified that experts agreed on only 60% of all classified cells ^26^. This can create challenges in identifying relevant cytomorphology-disease associations. Computational methods, which have shown promise in the characterization and prediction of MDS and AML using bone marrow slides ^27–29^ and identification of abnormal leukocytes ^30^, can help address these problems.

Here we present Haemorasis, a machine-learning protocol capable of automatically detecting and characterizing thousands of cells in PBS and apply it to a cohort of individuals with MDS and anemia, showing how it can be used to predict diseases and derive novel “morphotypes” — associations between cellular morphology and conditions. Particularly, we show that *SF3B1*-mutant MDS can be distinguished from other SF-MDS using cytomorphology and blood counts alone with high predictive performance, showing that hypolobulated neutrophils and large RBC are more prevalent in this MDS subtype. Additionally, using expert-annotated WBC and RBC, we show that virtual cell types are enriched for commonly recognized WBC and RBC types/abnormalities. Finally, we externally validate our approach, showing that it largely generalizes to other centers and WBS scanners.

## 2 Methods

### 2.1 Collection and digitalization of peripheral blood slides

Three retrospective sets of PBS from two different centers were digitized using two different slide scanners:

- **Training/discovery**
  - **CUH1:** used for training of cell detection. 54 PBS from randomly selected cases were used to develop the blood cell detection models described below. The PBS were automatically prepared using a Siemens Hematek system at Cambridge University Hospitals (CUH) and digitized using a Hamamatsu Nanozoomer 2.0 scanner.
  - **MLL:** used for training and discovery of disease-associated morphologies. 362 PBS from individuals with MDS with mutations in either *SF3B1, SRSF2, U2AF1* or *RUNX1*, iron deficiency anemia, megaloblastic anemia and controls. This cohort was used to develop the computational algorithms for predicting different clinical conditions and to train the quality control prediction model detailed below. The PBS were manually prepared in the Munich Leukaemia Laboratory (MLL) in Munich, Germany, and were digitized using a Hamamatsu Nanozoomer 2.0 scanner. Coverslips were added specifically for the digitalization as the routine cytomorphological characterization performed by experts at the MLL is performed using oil immersion microscopy and does typically not require coverslips.
- **Validation – CUH2:** used for validation of disease predictions. 68 PBS from individuals with MDS with mutations in either *SF3B1* or *SRSF2*, iron deficiency anemia, megaloblastic anemia and hematological normal controls. The PBS were prepared at Cambridge University Hospitals, UK, and digitized using an Aperio AT2. Slides belonging to individuals with MDS were prepared manually, whereas the rest were prepared automatically using a Siemens Hematek system.

Each slide was inspected individually. Inadequately prepared (excessive amount of stain) or scanned slides (the whole slide was blurred) were excluded from further analysis. Consequently, 11 slides from the MLL cohort and 1 slide from CUH2 were removed. Finally, for external validation, we excluded 4 PBS in CUH2 since they belonged to the same individual, keeping only one slide per individual. The condition-specific composition of each cohort, as well as the center from which they were retrieved and the slide scanner used, is detailed in **Table 1**. Characterizations of the individuals in the MLL cohort regarding age, sex, blood counts and clinical diagnostic and of the individuals in CUH2 regarding blood counts and clinical diagnostic are available in **Supplementary Table S1** and **Supplementary Table S2**, respectively.

**Table 1:**
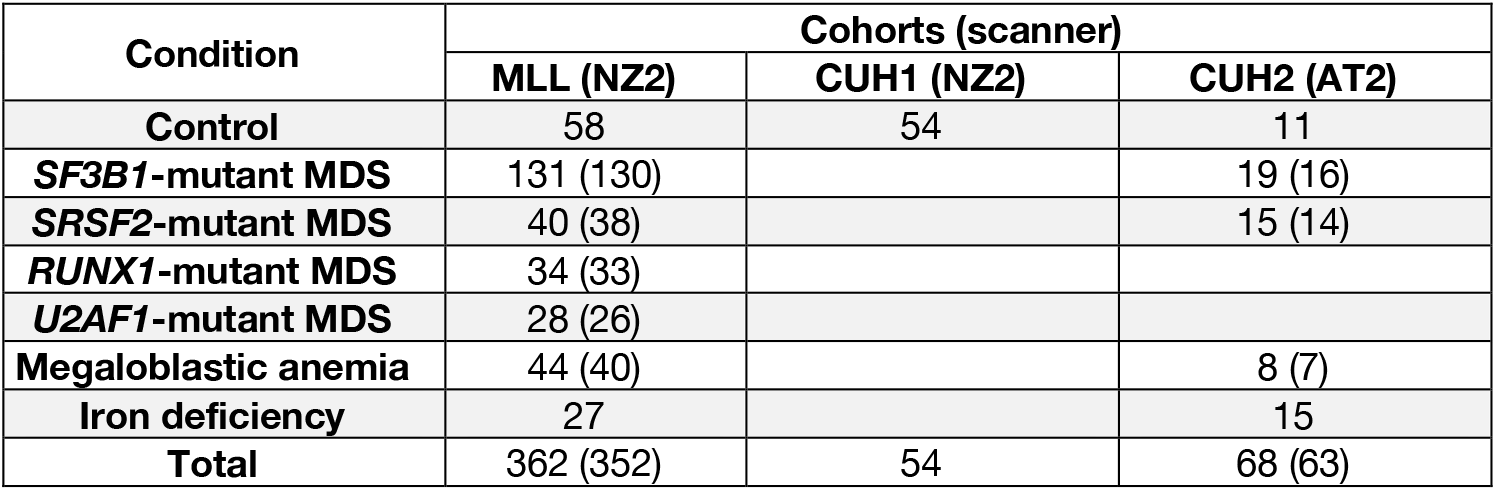
Condition-specific composition of each cohort. Numbers in brackets represent the total after excluding poor quality digitized PBS. Cohorts were retrieved from the Munich Leukaemia Laboratory (MLL) or from Cambridge University Hospitals (CUH) using either a Hamamatsu Nanozoomer 2.0 (NZ2) or an Aperio AT2 (AT2).

| Condition | Cohorts (scanner) |  |  |
| --- | --- | --- | --- |
|  | MLL (NZ2) | CUH1 (NZ2) | CUH2 (AT2) |
| **Control** | 58 | 54 | 11 |
| ***SF3B1*-mutant MDS** | 131 (130) |  | 19 (16) |
| ***SRSF2*-mutant MDS** | 40 (38) |  | 15 (14) |
| ***RUNX1*-mutant MDS** | 34 (33) |  |  |
| ***U2AF1*-mutant MDS** | 28 (26) |  |  |
| **Megaloblastic anemia** | 44 (40) |  | 8 (7) |
| **Iron deficiency** | 27 |  | 15 |
| **Total** | 362 (352) | 54 | 68 (63) |

### 2.2 Haemorasis — computational detection and characterization of blood cells

Our blood cell detection and analysis pipeline — **Haemorasis** — focuses on analyzing PBS tiles (small, computationally tractable parts of the image; **Supplementary Methods**) and comprehends two steps:

1. Extraction of white and red blood cells and their morphological characteristics from PBS scans.
2. Discovery of disease associated morphotypes.

The first step entails i) PBS quality control, ii) RBC detection, iii) WBC detection, iv) cell characterization and v) cell annotation.

#### Quality control of slide tiles

To detect tiles of poor quality, a deep neural network based on DenseNet121 ^31^, selected for its fast inference and good predictive performance, was trained on 10,000 randomly selected 512×512 pixel images from 93 slides from the MLLS split into training (8,050 tiles from 75 PBS) and testing (1,950 tiles from 18 PBS). Each tile was labeled as “good quality” (or 1, n=2,225) or “poor quality” (or 0, n=7,775). “Poor quality” tiles were defined as tiles where the concentration of cells was too small (very few/no cells) or too large (high frequency of overlapping cells) or images were blurred (**Supplementary Figure S1**). We use the ImageNet-pretrained Densely Connected Convolutional Network DenseNet121, a deep-learning model previously trained on a very large dataset of diverse images, which we fine-tuned on our training and then tested on our non-overlapping testing set. By using a pre-trained model, we guarantee that our model already captures visual features important for human perception of what constitutes a good quality image. Tiles were artificially augmented (computationally altered) during training through random flips/rotations/JPEG compression. Training was performed using the Adam optimizer ^32^ for 25 epochs with a learning rate of 0.00005 (additional details in **Supplementary Methods**). The evaluation of this model was performed using its validation AUC, precision and recall (**Supplementary Methods**)

#### Red blood cell detection

The initial segmentation and detection of RBC candidates was performed using Algorithm 1, a simple algorithm based on morphological operations (**Supplementary Methods**). In short, edges and contours were detected using a Canny edge detector ^33^ and filtered appropriately to ensure that other objects (platelet clumps, groups of RBCs or individual WBCs) are not detected as RBCs (**Supplementary Methods**). This ensures that only RBC candidates whose morphology is not affected by its neighboring cells are detected/extracted.

After detection by Algorithm 1, RBC candidates are characterized (see “Characterization of blood cells” in Methods) and filtered with an extreme tree gradient boosting (XGBoost) model ^34^ capable of predicting which candidates are actual RBC (vs. “not RBC”). The XGBoost algorithm was selected for this task due to its fast implementation and excellent performance. To train this model, we picked a random set of 158 2096 × 2096 pixel tiles from CUH1 and split them into training (109) and testing tiles (48) and detected RBC using Algorithm 1. We then annotated 2262 RBC candidates for training (“RBC”: 1870; “not RBC”: 392) and 1107 (“RBC”: 962; “not RBC”: 145) for testing. We picked a maximum number of 50 weak estimators and DART boosting ^35^ for training and validated the model on the testing set. Given the relatively high class imbalance (6 of 7 objects belonged to the “RBC” class, such that calling all objects a RBC would still constitute good performance), the objective function (importance) of each “RBC” was scaled by a factor of 0.1 during training. The evaluation of this model was performed using its validation accuracy and *F*_1_ (**Supplementary Methods**).

#### White blood cell detection

Our WBC segmentation and detection algorithm is based on U-Net, a deep neural network developed for biological image segmentation ^36^. This model requires that all images of cells are annotated at the pixel level. To this effect, we labeled 158 2096 × 2096 pixel images from CUH1, comprising a total of 2,853 annotated WBC including a of which 2,747 were completely present in the center of the image and thus defined to be positively identified. Two additional images from MLLS and 30 images from CUH2 (containing 60 and 69 WBC, respectively) were annotated to test the generalization of our U-Net model. Different architectures were tested to assess whether network depth (the number of channels in each convolutional layer) had an impact on output; for this, different instances of a U-Net model were trained where the depth of each layer was multiplied by 1.0, 0.5 and 0.25 and rounded to the nearest integer (this varies the number of features that a model seeks to identify and use to explain the data). We train each U-Net model on 512 × 512 pixel tiles obtained from the original 2096 × 2096 pixel images with a learning rate of 0.001 and a weight decay of 0.005 with a weighted cross-entropy loss using the Adam optimizer ^32^, whilst making extensive use of data augmentation to ensure better generalization to other images/datasets (**Supplementary Methods**). We used the Jaccard index (otherwise known as “intersection over union” or IoU; **Supplementary Methods**) to assess the performance of these models and tested whether test-time augmentation (TTA) ^37^ and prediction post-processing (**Supplementary Methods**) had a positive impact on performance.

Finally, we segmented the nuclei of WBCs. As nuclei stain darker than the rest of each WBC, we clustered the pixels on each segmented WBC using k-means clustering (k=2) into two separate clusters as described before ^38^. This enables easy assignment of each pixel to the darkest (nucleus) or brightest (cytoplasm) parts of the cell.

#### Morphometric characterization

After detecting and segmenting each cell, we calculate a set of cytomorphological descriptors for each one. The descriptors used here were implemented in a custom Python script, but the large majority are also present in widely used bio-image analysis software programs ^39,40^ or described in publications reviewing morphometry in image analysis ^41^. We characterize every RBC and WBC using the features described in **Supplementary Table S4**. Additionally, we characterize the nucleus of each WBC using a reduced set of features, also detailed in **Supplementary Table S4**. Overall, we capture 53 features for each WBC (42 for the cellular characterization and 11 for the nuclear characterization) and 42 features for each RBC.

To account for the different resolutions (0.2268 micrometers/pixel for Hamamatsu NanoZoomer 2.0; 0.2517 micrometers/pixel for Aperio AT2) we rescale the images in CUH2 by a factor of 1.1098 before characterizing the cells to ensure these features are comparable.

#### Morphometric moments

As morphometric moments, the parametric characterizations of the distribution of each feature within each PBS, we calculate the mean and variance of each RBC and WBC feature on each PBS. Using this definition, we describe the cytomorphological landscape of each PBS as a set of 190 features ((42 + 53) × 20).

### 2.3 Cytomorphological prediction of clinical conditions

We assess the utility of our blood cell detection and characterization protocol by focusing on four distinct binary prediction tasks:

1. Disease detection — identifying the presence of either deficiency anemia or MDS vs. normal blood;
2. Disease classification — distinguishing between deficiency anemia and MDS;
3. MDS genetic subtyping: distinguishing between *SF3B1*-mutant and *SF3B1*-wildtype (*SF3B1*-wt) MDS;
4. Anemia classification — distinguishing between iron deficiency anemia and megaloblastic anemia.

#### Machine-learning using morphometric moments

We assess the predictive performance of morphometric moments using elastic net regression (glmnet) ^42^ and 5-fold cross-validation on the MLLS, calculating the cross-validated area under curve (AUC) of the receiver operating characteristics curve (ROC) by concatenating all cross-validated predictions ^43^. We also test whether using blood counts (WBC counts (WBCC; cells/*µ*L), hemoglobin concentration (Hb; g/dL) and platelet counts (Plt; platelets/*µ*L)) as predictors improves classification performance, and preprocess features to avoid large numerical differences. Finally, we assess how different features and groups of features contribute to prediction (**Supplementary Methods**).

#### Morphotype analysis

Typically, when diagnosing hematological conditions from a PBS, an expert focuses on identifying abnormal cell types and quantifying their relative prevalence ^2^. In more abstract terms, the expert has to classify objects in a set in order to quantify the prevalence of abnormalities, and classify the set according to the prevalence of different classes of abnormalities. This is essentially a problem of multiple instance learning (MIL), a machine-learning field that focuses on classifying a set/group of objects based on its composition. Some MIL methods combine this group classification task with the auxiliary task of learning a function that maps objects to a vocabulary (set of terms) and uses the presence or frequency of instances in each term in the vocabulary to predict a classification for the group ^44^.

The use of this abstraction can help devise an approach that does not only classify PBS into binary classes (i.e. “normal” vs. “disease”), but also can classify each individual object into specific categories that enhance the classification process. In our case, we are interested in classifying cells (objects) into automatically defined and clinically-relevant categories which we call computational morphotypes (CM) — and use the prevalence of these CM to predict conditions from PBS. For this, we use Morphotype analysis, a method to (i) identify relevant morphological classes of cells (CMs) without recourse to human-based classification and (ii) distinguish between conditions using morphotype proportions, which can also incorporate other data types such as blood counts. This method considers WBC and RBC individually and classifies each cell into a given CM, using the average prevalence of CM to predict conditions. Morphotype analysis is fully differentiable and, as such, we optimize it using gradient-descent based methods, particularly Adam ^32^. We test Morphotype analysis performs with a single set of CM to predict all four tasks specified above (multiple objectives — MO) and with a different model for each task (single objective — SO), and how different assumptions regarding the number of CM (25 and 50 for MO and 10, 25 and 50 for SO) impact predictions. For inference, we also consider how using only stable CM — CM which are consistently detected across different folds — can improve the results by eliminating biased and fold-specific CM. More details on Morphotype analysis are available in the **Supplementary Methods**.

### 2.4 Validation

#### Expert annotation of blood cells

Three expert hematologists annotated 1,746 RBC and 1,600 WBC automatically detected in MLLS to assess whether CMs were enriched in any expert-annotated blood cell type. RBC were expert-annotated into the following classes: “normal”, “nucleated RBC”, “spherocyte”, “target cell”, “irregularly contracted RBC”, “dacrocyte”, “keratocyte”, “echinocyte”, “eliptocyte” and “acanthocyte”, or into: “incomplete RBC”, “multiple RBC”, “no RBC”, “platelet/dye”, “nearby platelet” and “unable to tell”. WBC were annotated with the following classes: “neutrophil”, “basophil”, “eosinophil”, “band cell”, “lymphocyte”, “monocyte”, “blast”, with the artefacts “incomplete WBC”, “multiple WBC”, “no WBC”, “poor resolution”, “RBC”, “nucleated RBC”, “reticulocyte”, “fragmented/unrecognizable” and “platelet clump”. Enrichment for each CM was calculated as the ratio between the proportion of cells of a given expert-annotated cell type belonging to that CM (type/CM), divided by the proportion of cells of the given cell type in the entire set of expert-annotated cells (type/total).

#### External validation

To externally validate the performance of our predictive methodologies — glmnet with morphometric moments and Morphotype analysis — we use CUH2, a cohort of 63 PBS whose composition is similar to the MLL training cohort. We test all the best performing models on this cohort, reporting their AUC estimates and confidence intervals (calculated as ^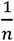^, where *n* is the size of the validation sample).

### 2.5 Statistical analysis

All statistical analyses in this work were conducted using the R statistical software (v3.6.3) ^45^. Machine-learning models were implemented in either R for the elastic net model with the glmnet package ^42^ or Python for morphotype analyses with PyTorch ^46^.

### 2.6 Data availability

The different cohorts of digitized peripheral blood slides can be made available upon request. The annotated datasets for tile quality classification, white blood cell segmentation and red blood cell filtering can be found at https://doi.org/10.6084/m9.figshare.19153760. The machine-learning model parameters are available at https://doi.org/10.6084/m9.figshare.19164209. The necessary data to run Morphotype analysis is available at https://doi.org/10.6084/m9.figshare.19372292. The output of the Morphotype analysis, as well as the expert annotated cells, and the data necessary for downstream analysis are available at https://doi.org/10.6084/m9.figshare.19369391 and https://doi.org/10.6084/m9.figshare.19371008, respectively.

### 2.7 Code availability

We have made the Haemorasis pipeline available in https://github.com/josegcpa/haemorasis and as a Docker container in https://hub.docker.com/repository/docker/josegcpa/blood-cell-detection (instructions to run this are provided in **Supplementary Methods**). The code necessary to run Morphotype analysis (mil-comori) and the statistical analysis and plot generation (analysis-plotting) for this paper are available in https://github.com/josegcpa/wbs-prediction. The code necessary for training, testing and inference using the quality control network is available in https://github.com/josegcpa/quality-net. The code necessary for training, testing and inference using the U-Net is available in https://github.com/josegcpa/u-net-tf2.

## 3 Results

### 3.1 The MLL cohort captures previously described clinical features of MDS and anemia

We start by confirming that the MLL cohort is a representative set of PBSs. Individuals with MDS were older than the rest of the MLL cohort with a bias towards males as previously reported ^47^ — indeed, the chance of having MDS in our cohort increased by 12% every year, with males being more than twice as likely to have MDS (*p* = 8 * 10−16 and *p* = 0.00017, respectively, for the binomial regression of MDS diagnosis based on age and sex; **Figure 1a,b**).

**Figure 1.**
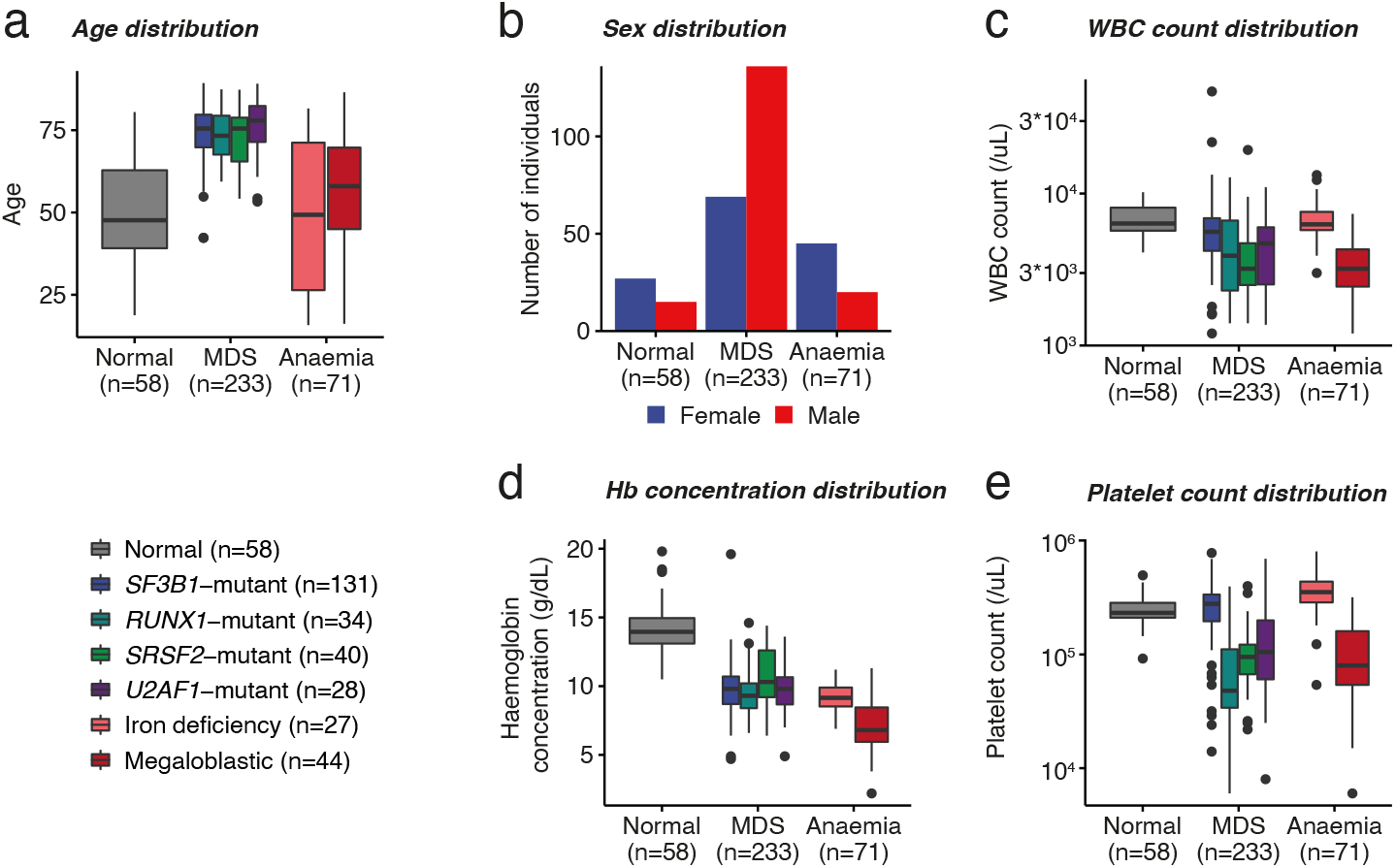
General features of anemias and myelodysplastic syndromes (MDS) in the MLL cohort. **a,b** — Age and sex distributions of individuals according to different conditions, respectively. **c,d,e** — White blood cell (WBC) counts, hemoglobin concentration and platelet counts, respectively, according to different conditions.

Additionally, MDS and deficiency anemias were associated with leukopenia (1,200 (*p* = 0.04) and 1,800 (*p* = 0.009) fewer WBC/*µ*Lin these conditions, respectively, for the linear regression WBC counts against binary MDS and anemia diagnosis indicators; **Figure 1c**). However, this leukopenic tendency in anemias is driven by megaloblastic anemia — indeed, whereas iron deficiency is indistinguishable from controls, megaloblastic anemia has approximately 3,200 fewer WBC/µ*L* than controls (*p* = 6 × 10^−14^ for a two sample t-test) in keeping with previous studies ^10,48,49^. Hb are also much lower in MDS and anemias (**Figure 1d**) — indeed, the Hb of these individuals was lower than that of normal individuals by 4.34 and 6.38 g/dL, respectively (*p* < 2 × 10^−14^ and *p* < 2 × 10^−14^, respectively, for the linear regression Hb against binary MDS and anemia diagnosis indicators). No detectable difference between normal individuals and individuals with either MDS or anemia is observable when it comes to platelet counts (Plt), but megaloblastic anemia have approximately an expected 146,000 fewer platelets/*µ*L than controls (*p* = 3 × 10^−12^ for two sample t-test; **Figure 1e**) as previously reported ^48^.

Finally, we also observed that *SF3B1*-mutant MDS displayed distinct features compared to *SF3B1*-wt MDS — particularly, WBC and Plt are comparable to those of controls and higher than those found in *SF3B1*-wt MDS (*p* = 0.3 and *p* = 0.15 for two sample t-tests comparing WBC and Plt between *SF3B1*-mutant MDS and controls; *p* = 0.002 and *p* < 2 × 10^−14^ for two sample t-tests comparing WBC and platelet counts, respectively, between *SF3B1*-mutant and *SF3B1*-wt MDS), in keeping with previous reports ^15,16^.

### 3.2 Computational cytomorphology of peripheral blood slides

We detect cells in PBS using the Haemorasis pipeline outlined in **Figure 2a**. For the first stage of this method, quality control of PBS tiles, we trained a DenseNet121, a small and fast neural network with good performance ^31^, to predict whether specific tiles are of “good” or “poor” quality (**Supplementary Figure S1a**). This (i) reduces the chance of falsely detecting cells in the downstream analysis (false positives) by reducing variation associated with preparation or digitalization in the tiles and (ii) limits processing to the interpretable part of the PBS (usually less than 20% of the total area). To train this model, we manually annotated a dataset from the MLL cohort PBSs composed of 8,050 tiles as being of “good” or “poor” quality, and validated the trained model on a smaller dataset of 1,950 tiles from different the MLL cohort PBSs. Overall the model performed well and comfortably within what we required for this filtering step (validation *AUC* = 93.4%, *recall* = 82.2% and *precision* = 85.2%). Of note, most errors were not clear mistakes, but usually represented tiles of borderline quality, with the predicted “good” quality regions corresponding to the monolayer (**Supplementary Figure S1b**).

**Figure 2.**
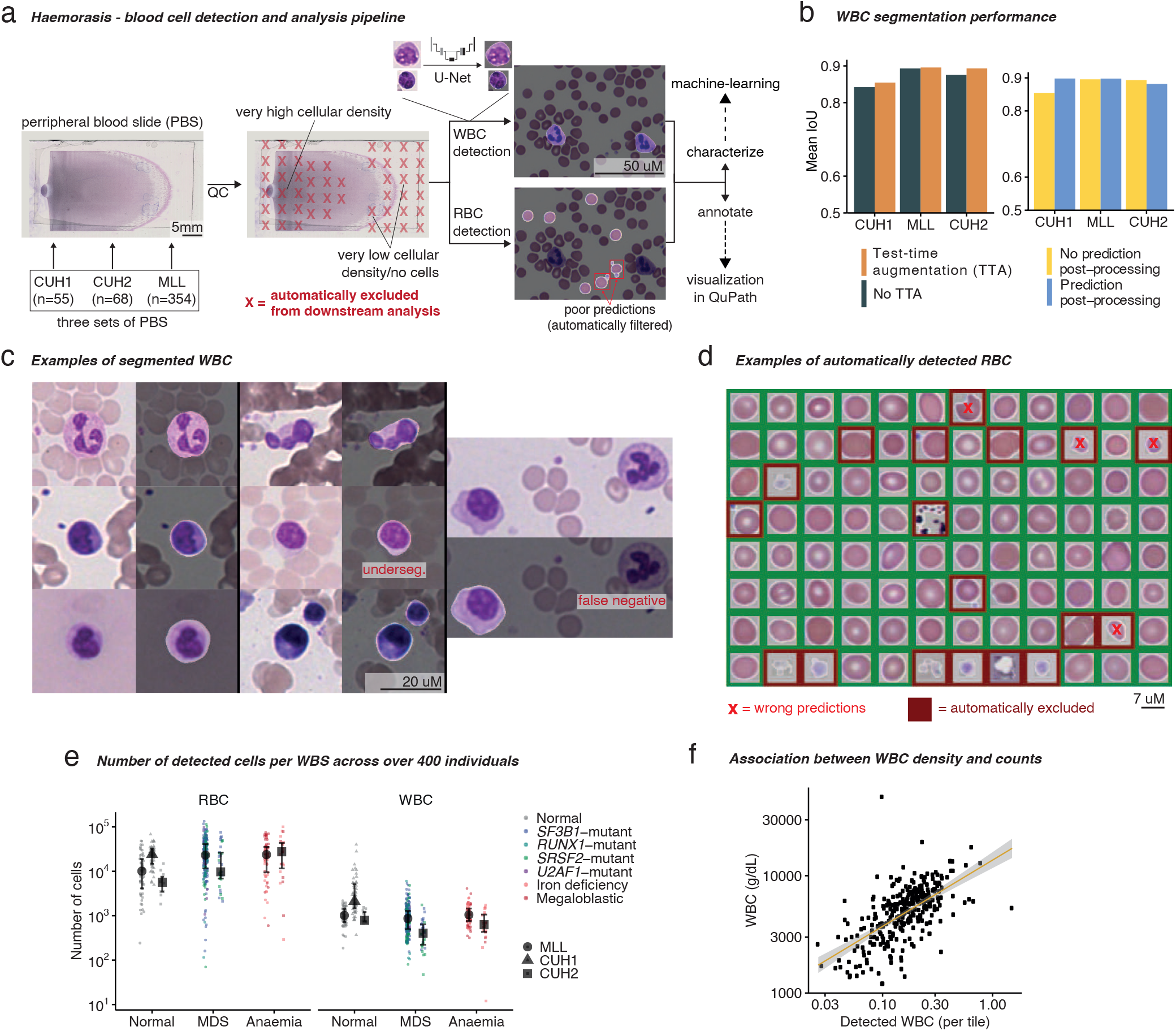
Haemorasis – automated detection and analysis of blood cells in peripheral blood slides (PBS). **a** — Haemorasis: detection and characterization of blood cells in PBS using computer vision and machine-learning. First, the digitized PBS goes through a deep-learning-based quality control (QC) algorithm that filters out parts of the PBS which are too high or low in cellular density or too blurred. Then, white blood cells (WBC) and red blood cells (RBC) are detected separately — WBC are detected using a U-Net model, a deep-learning algorithm for segmentation, while RBC are detected using simple computer vision methods and filtered using machine-learning. Following this, each individual cell is characterized in terms of shape, texture and color distribution and annotated for visualization in QuPath ^50^. **b** — U-Net performance on WBC segmentation. Segmentation post-processing was tested on test-time augmented images. **c** — WBC detection examples and possible errors (underseg. = image undersegmentation error). **d** — RBC detection examples and examples of wrongly detected and filtered RBC. Here, all RBC were detected using a simple computer vision protocol, but wrong detections were filtered out using machine-learning (red background). **e** — Number of detected blood cells stratified by clinical classification. **f** — Association between number of detected WBC using our protocol and WBC counts (robust *R*^2^ = 0.39, *CI*_95%_ = [0.30,0.49]).

Next, we detect both WBC and RBC on “good” quality tiles. To detect WBC, we trained a deep neural network based on U-Net ^36^ with extensive data augmentation (random alterations to the image which make the algorithm more robust) during training (**Supplementary Table S3**) on a dataset with >2,800 manually annotated WBC in PBS from CUH1 and validated this on separate testing sets from CUH1, CUH2 and the MLL cohort, confirming the good performance of the model through visual inspection (**Figure 2b,c, Supplementary Figure S2**). A smaller version of the U-Net with fewer parameters was the best performing model for WBC segmentation, possibly by decreasing the chance of overfitting, with prediction post-processing and test time augmentation (TTA) improving predictions (**Figure 2b, Supplementary Figure S2**). While some errors were detected upon visual inspection of segmented images (**Figure 2c**), the model performed well on data from different cohorts (average intersection between ground truth and prediction over the union of both (IoU) of 84.4% for CUH1 and 88.9% for CUH2 and the MLL cohort; **Figure 2b**), indicating these errors are small and rare. RBC detection was performed using a simple computational protocol with predictions filtered using XGBoost, a fast and scalable machine-learning algorithm ^34^ (**Figure 2d**). Filtering ensured that non-RBC objects in PBS were removed and was validated on an independent test set, showing high discriminative ability with validation *accuracy* = 90.6% and *F*_1_ = 94.2% (*F*_1_ is a better metric for imbalanced data), reducing the estimated rate of false positives from 1 in 7 to 1 in 300.

Across all cohorts, for each PBS we are able to detect an average of 26,000 (range 70 to 133,916) RBC per PBS (a total of 12,042,425 RBC) and around 1,400 (range 12 to 39,862) WBC/PBS (a total of 646,952; **Figure 2e**)). We show that the cellular density of detected WBC in the PBS correlated with WBCCs from automated analysers, validating our detection protocol through an orthogonal approach (robust *R*^2^ = 0.39, *CI*_95%_ = [0.30,0.49]; **Figure 2f**). Finally, we characterized all individual cells using well-known morphological features used in other morphometric software programs ^39–41^ (**Supplementary Table S4**; **Supplementary Figure S3**). For each cell, we quantified its size, shape, color distribution and texture and for WBCs we also characterized their nuclear size and shape (**Supplementary Figure S3**).

### 3.3 Morphological heterogeneity informs disease prediction

We test four distinct tasks to determine whether our protocol for automated morphometric characterization can be used to meaningfully predict conditions from PBS: (i) disease detection (health vs. disease), (ii) disease classification (deficiency anemia vs. MDS), (iii) MDS genetic subtyping (*SF3B1*-mutant MDS vs. other MDS) and (iv) anemia classification (iron deficiency vs. megaloblastic anemia). We observed that morphometric feature distributions, represented by feature mean and variance across all cells in a given slide, differed across different conditions in the MLL cohort (**Supplementary Results**; **Supplementary Figure S4**). This qualitative assessment was corroborated by fitting a binomial elastic-net regression model (glmnet) ^42^ for each of the four tasks considered using morphometric moments (slide means and variances) in addition to WBC, Hb and Plt counts. Performance was evaluated using 5-fold cross validation, where the data is split into 5 non-overlapping validation sets while the rest is used to train the model, leading to less biased models ^51^.

Morphometric regression showed high cross-validated predictive performance across all four tasks (**Supplementary Figure S5a,b**), including an AUC of 89.7% for MDS genetic subtyping (**Supplementary Figure S5b**). We additionally show that blood counts are highly predictive of *SF3B1*-mutant MDS as indicated in **Figure 1c,e** and previous publications ^15,16^ (**Supplementary Figure S5b-c**). Of note, morphological feature variation (WBC and RBC variance) had a significant impact on prediction, revealing that cytomorphological heterogeneity is important for diagnosis (**Supplementary Figure S5c**), as previously suggested for red cell distribution width ^52^. Finally, the relative importance of different features revealed important trends in the detection of different conditions. For instance, *SF3B1*-mutant MDS was characterized by higher platelet counts, larger RBC and smaller WBC nuclear area (**Supplementary Figure S5d**). However useful, this protocol can make the retrieval of illustrative examples of blood cells more challenging, i.e. while the use of means reveals that larger RBC or more irregular WBC are associated with different conditions, variances do not lend themselves to illustrative explanations, making the demonstration of their importance more elusive.

### 3.4 Discovering diagnostically relevant morphotypes

At first inspection, the distribution of cytomorphological characteristics of different conditions revealed an interwoven landscape without immediately recognizable cell clusters (**Figure 3a**). However, it becomes apparent that different parts of the cytomorphology space are differentially populated by different conditions. In order to partition this space and define cytomorphological phenotypes associated with specific conditions, we use a multiple instance learning approach that clusters cells based on their cytomorphological characteristics such that the resulting clusters – in the following referred to as computational morphotypes – are relevant to the aforementioned diagnostic tasks (**Figure 3b**; **Supplementary Methods**).

**Figure 3.**
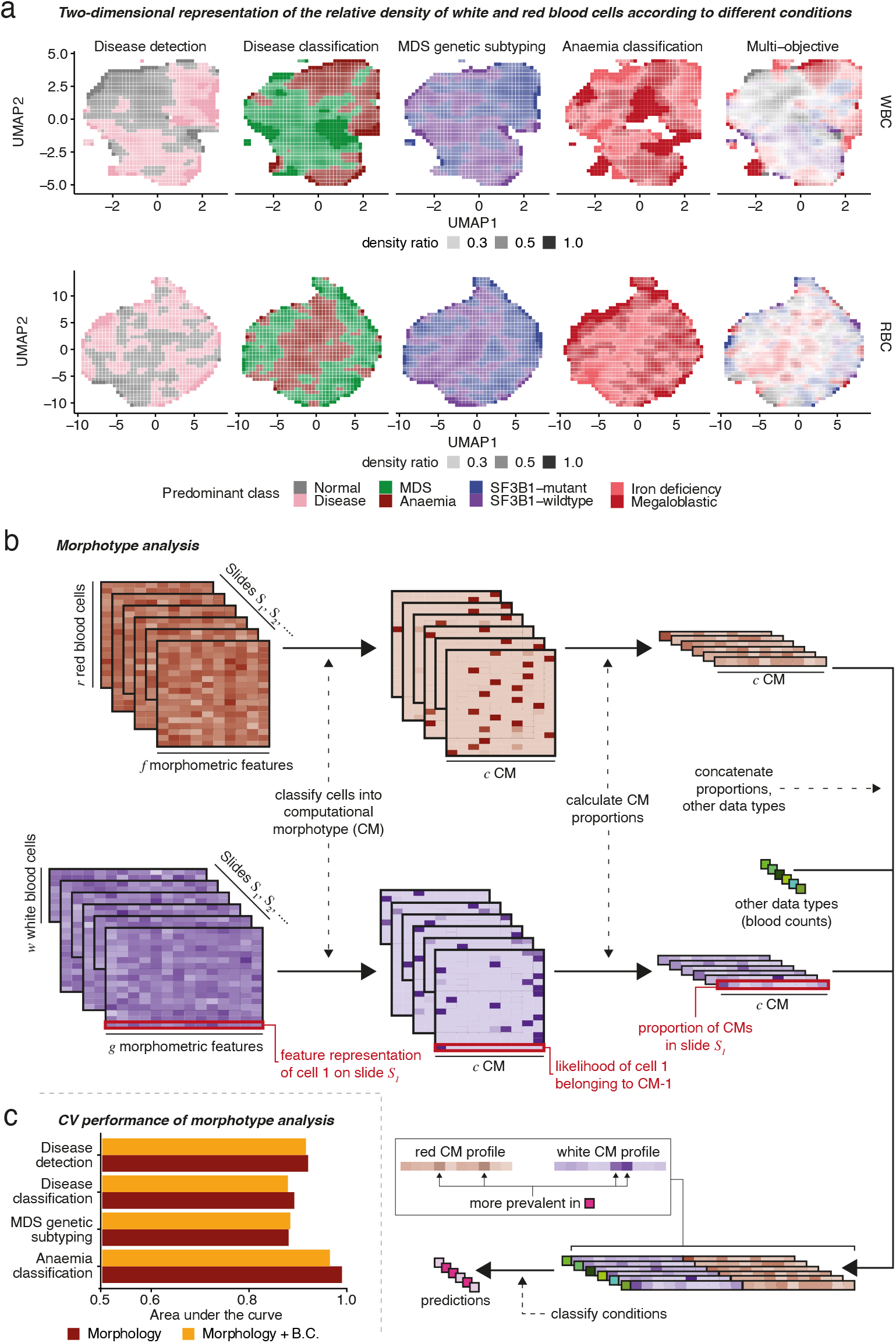
Morphotype analysis. **a** — Two-dimensional UMAP density ratios of the features of individual WBC (top) and RBC (top). Density ratios are calculated by dividing the local density for the predominant class by the sum of the local density of all classes. **b** — Schematic representation of Morphotype analysis. Individual WBC and RBC are clustered into computational morphotypes (CMs) and the proportions of each morphotype on each slide is used to predict different conditions. This allows us to identify which morphotypes are associated with different conditions. **c** — Performance of prediction using Morphotype analysis.

We performed morphotype analysis considering the four objectives described earlier and established stable morphotypes consistently found through 5-fold cross-validation (**Supplementary Methods**). Morphotype analysis performed similarly to glmnet with morphometric moments when predicting conditions (**Figure 3**; **Supplementary Figure S6, Supplementary Figure S7**), however with the added benefit of producing a small number of characteristic human-interpretable, disease-associated cytomorphologies. This approach revealed 8 stable WBC morphotypes (denoted WCM 1-8), accounting for 60% of WBCs in normal samples, as well as 12 stable RBC morphotypes (RCM 1-12) comprising 90% of RBCs (**Supplementary Figure S8**). These stable WBC and RBC morphotypes displayed distinct cytomorphological characteristics, whilst the remaining morphotypes were found to be of variable nature or were not associated with any of the conditions and thus not further analyzed. Among these stable morphotypes, 7 WCMs and 7 RCMs exhibited robust associations with specific clinical conditions discussed below (**Figure 4a-c, Supplementary Figure S9**).

**Figure 4.**
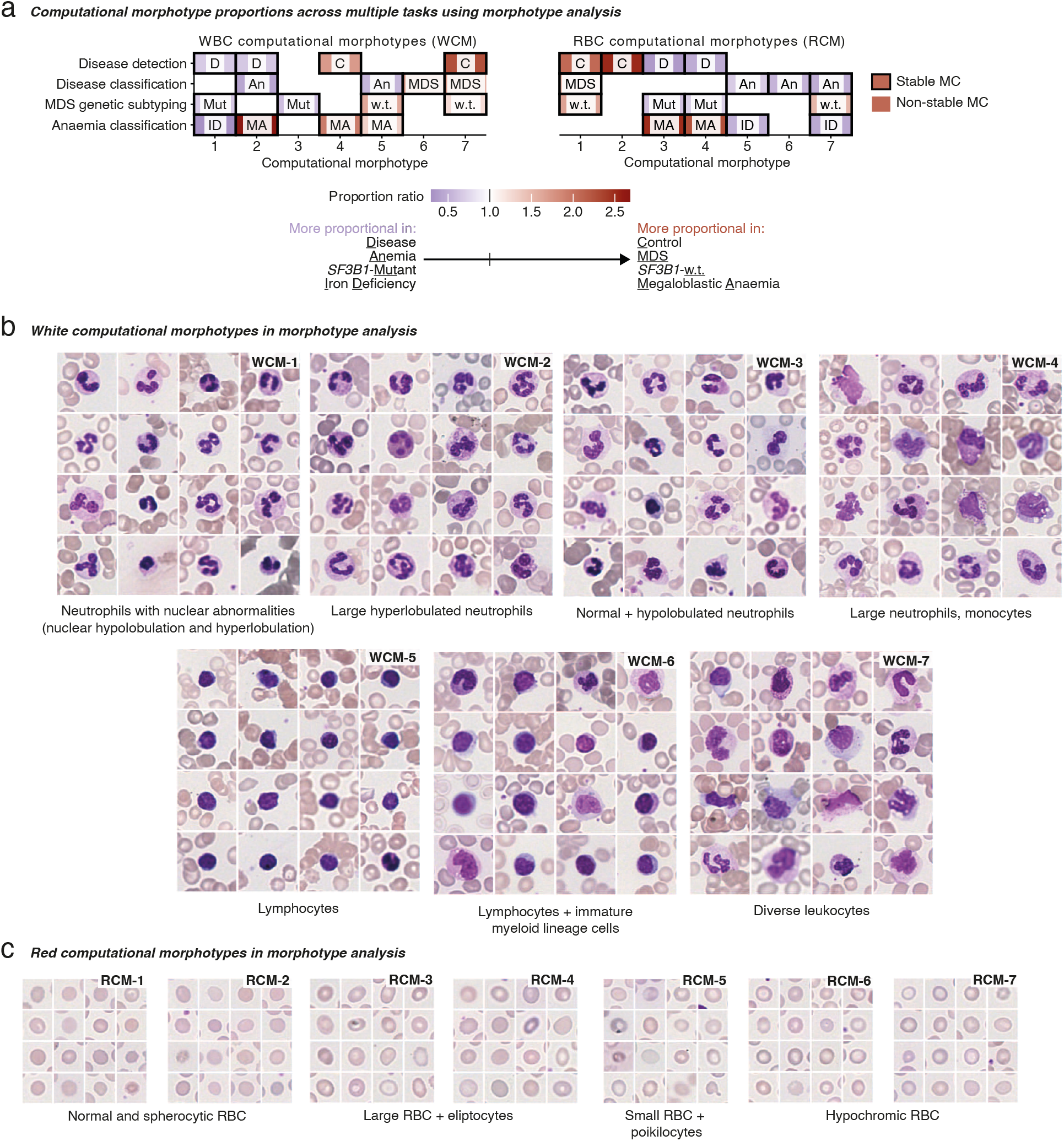
Computational morphotypes across conditions. **a** — The relevant prevalence of disease-associated computational morphotypes. For this heatmap, the 5 morphotypes with the highest absolute difference in median effect size between conditions are selected for each task, and proportion ratios were calculated as the ratio of the median proportion for each condition. **b** — White blood cell morphotypes (WCMs) for different conditions from Morphotype analysis. **c** — Red blood cell morphotypes (RCMs) for different conditions from Morphotype analysis. The labeling for WCMs in **b** and the WBC heatmap in **a** is the same, as well as the labeling for RCMs in **c** and the RBC heatmap in **a**.

Among the stable WBC morphotypes, four mostly consisted of different neutrophil morphologies (WCM-1,2,3 and 4 in **Figure 4a,b**), highlighting their cytomorphological diversity and diagnostic relevance. WCM-5 contained small lymphocytes, WCM-6 larger lymphocytes and myeloid progenitors, whereas WCM-7 consisted of diverse myeloid cells. We confirm clinically-relevant cellular phenotypes such as the increased prevalence of morphological anomalies in neutrophils in cases of MDS and deficiency anemia (WCM-1 and 2); in MDS we found a lower prevalence of lymphocytes (WCM-5) and increased prevalence of immature myeloid cells (WCM-6) as previously suggested ^53,54^. Morphotype analysis also identified novel clinically-relevant phenotypes — particularly, WCM-3 (normal hypolobulated neutrophils) appeared to be more prevalent in *SF3B1*-mutant MDS, and larger and/or hyperlobulated neutrophils were more prevalent in megaloblastic anemia than in iron deficiency (WCM-2 and 4). We confirm these using single-objective Morphotype analysis, where morphotypes are trained on a single task only (**Supplementary Figure S10**). Finally, we found that WCM-5 (small lymphocytes) were more prevalent in anemia when compared with MDS.

Stable RBC morphotypes showed more subtle differences. Some morphotypes were relatively more normal — RCM-1 and 2 contained mostly normal or spherocytic RBCs (**Figure 4a,c**) — whereas others (RCM-3 and 4) captured larger RBC and elliptocytes. RCM-5 captured relatively small RBC and some poikilocytes, and RCM-6 and 7 captured hypochromic RBCs. We show that RCM-6 and 7 (hypochromic RBCs) were more typical of anemia than MDS as previously reported in iron deficiencies ^1^, with RCM-7 being more prevalent in iron deficiency compared to megaloblastic anemia. In *SF3B1*-mutant MDS, RCM-3 and 4 (large RBC and elliptocytes) were more prevalent compared with other MDS, while RCM-5 (poikilocytic RBC) were more common in iron deficiency compared to megaloblastic anemia.

Of note, morphotype frequency offers more intuitive and tangible explanations for the associations of morphometric moments with certain diagnoses described in the previous section. If, on average, there is a higher prevalence of morphotypes with specific morphometric characteristics in a certain condition, this will manifest as a relation with shifts in the mean of a feature. Similarly, it may be that multiple morphotypes change in prevalence such that the morphological diversity changes, manifesting in an association between the morphometric variance and a condition (**Figure 5, Supplementary Figure S11**). For example, the variance of the WBC nuclear perimeter, shown to be associated with in disease detection (**Supplementary Figure S5**) can be explained by the differential frequencies of different WCMs (**Figure 5a**): the higher prevalence of WCM-7 drives the increased heterogeneity this feature in normal individuals. In disease classification, we can further observe how the increased variance of RBC shape irregularity (standard deviation of the centroid distance function) in anemia compared to MDS is partly explained by the elevated prevalence of RCM-5, 6 and 7 (relatively circular, some poikilocytes) and lower prevalence of RCM-3 and 4 (larger and more elliptic; **Figure 5b**). Finally, WBC nuclear convexity exhibits a stronger bimodality and therefore greater variance in *SF3B1* mutant MDS cases, driven in part by WCM-1 and 3 (**Figure 5c**). Also, there is a clear increase in the mean of RBC area in *SF3B1*-mutant MDS, due to the higher prevalence of RCM-3 and 4 and lower prevalence of RCM-1 and 7 in *SF3B1*-mutant MDS (**Figure 5d**).

**Figure 5.**
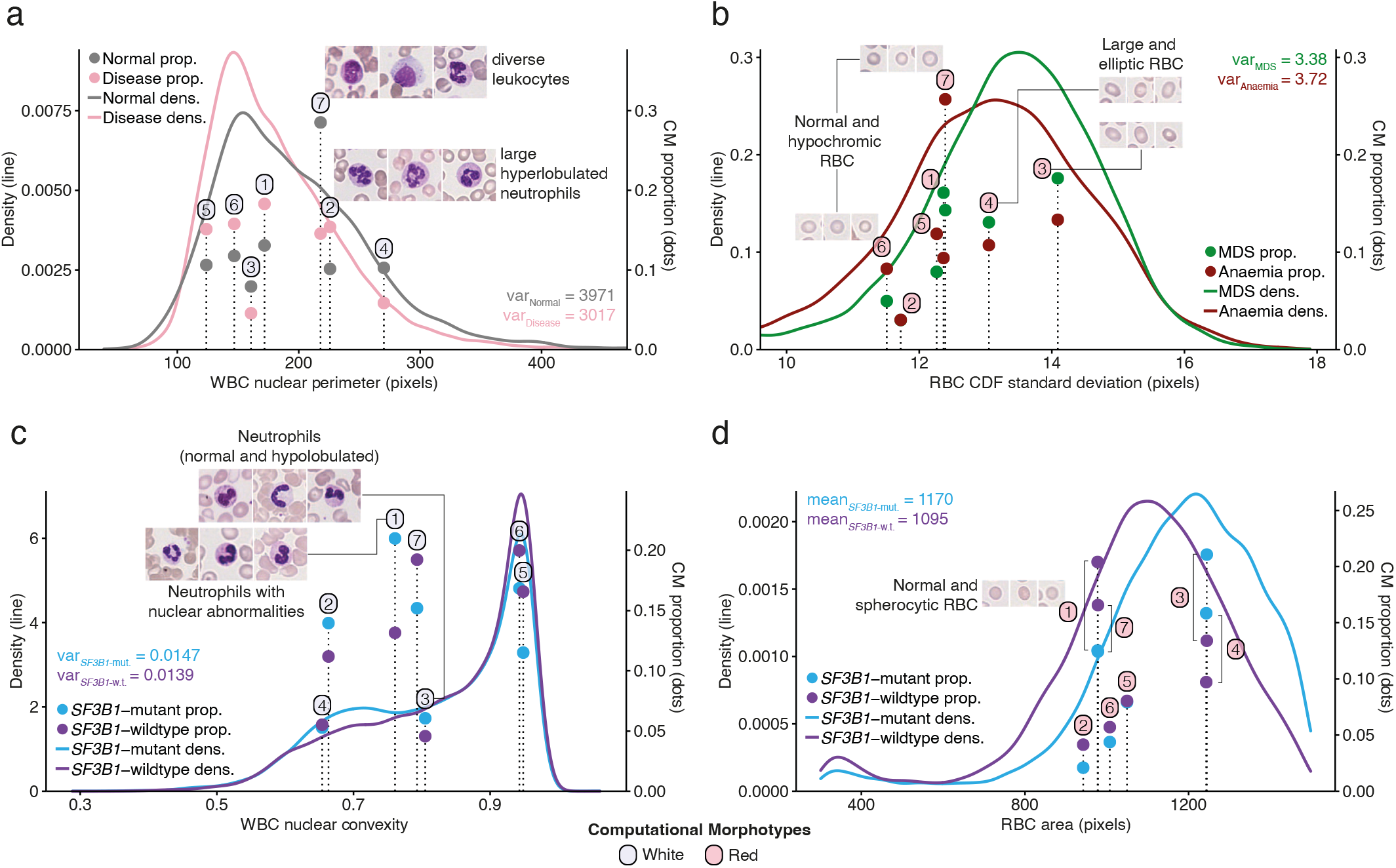
Computational morphotype proportions explain morphometric feature distributions. **a-c** — Examples of the relationship between condition-specific WCM (left column, a,c) and RCM (right column, b,d) proportions and differences in morphometric feature variances for disease detection (WBC nuclear perimeter), disease classification (RBC standard deviation of the centroid distance function (CDF)) and MDS genetic subtyping (WBC nuclear convexity), respectively. **d** — Example of the relationship between condition-specific morphotype proportions and differences in RBC area mean for MDS genetic subtyping. Morphometric feature distributions and stable morphotype classifications were derived using a subset of 31,119 RBC and 31,884 WBC from the MLL cohort. The positions of the bars corresponding to different morphotypes are calculated as the mean feature value for each morphotype. Each label corresponds to the nearest bar top, and only morphotypes presented in **Figure 5** are annotated.

### 3.5 Computational cytomorphology validation

To confirm the nature of the computational morphotypes and their diagnostic associations, we performed: (i) a blinded annotation of cell types by expert hematologists and (ii) a validation of their predictive value in the CUH2 cohort. First, we assessed whether the morphotypes determined by Morphotype analysis were enriched in known cell types. Three hematologists labeled up to 1,746 RBC and 1,600 WBC. This demonstrates that morphotypes are enriched in known RBC and WBC types (**Figure 6a,b**), but concordance was limited for some rare cell types (particularly hypolobulated neutrophils and blasts; **Supplementary Figure S12**). These results are also observed in single objective morphotype analysis, particularly for disease detection and classification models (**Supplementary Figure S13**). Furthermore, the CMs enriched in artifacts were rarely enriched with known cell types.

**Figure 6.**
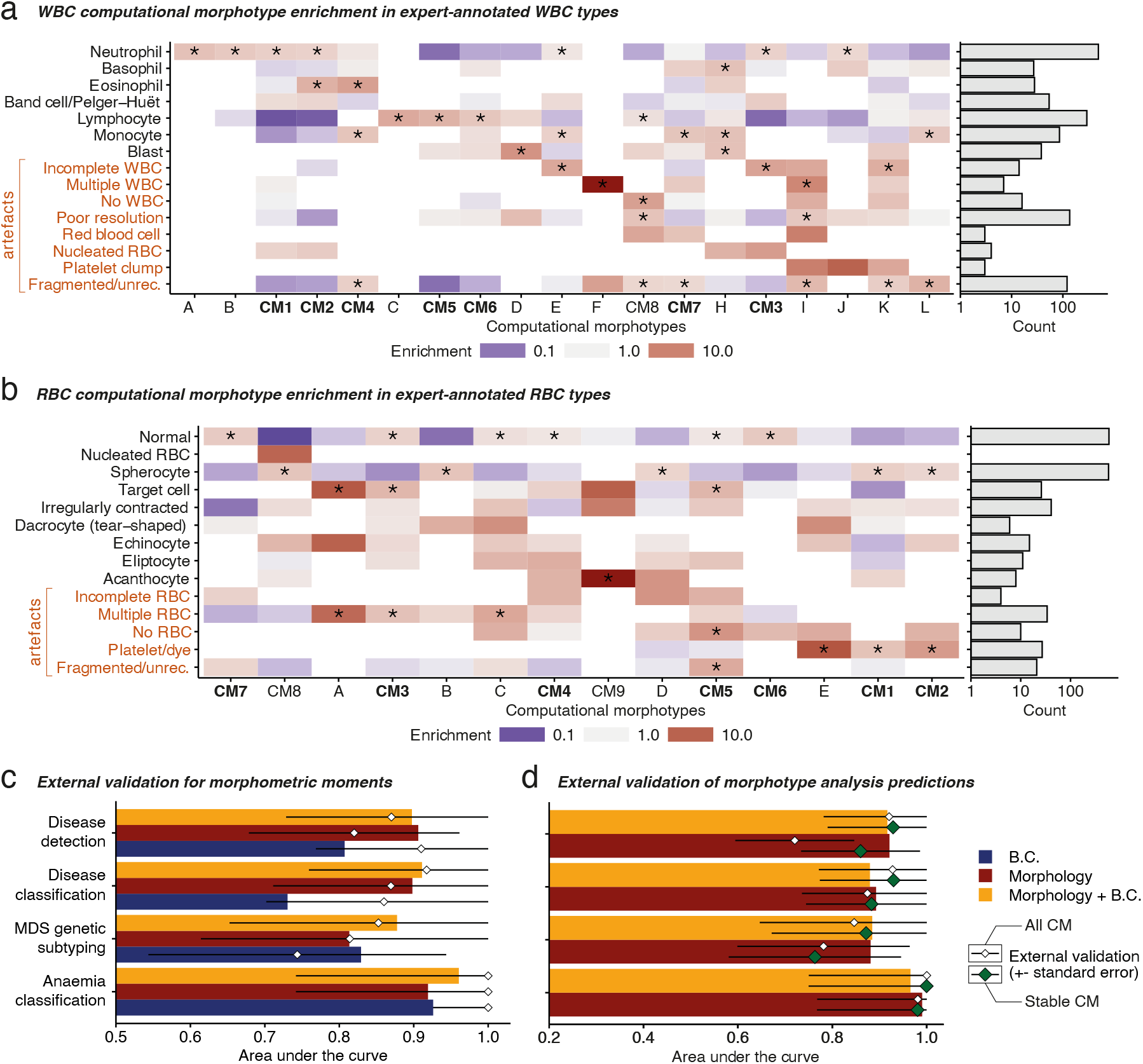
Expert and external validation of computational cytomorphology. **a** — Correspondence between WCMs and expert annotated WBC types (left) and number of annotated WBC types. **b** — Correspondence between RCMs and expert-annotated RBC types (left) and number of annotated RBC types. **c** — External validation for glmnet. **d** — External validation performance for the Morphotype analysis with all morphotypes (small white diamonds) and using only stable morphotypes (green diamonds). Enrichment values in **a** and **b** marked with an asterisk “*” are significant for a chi-squared test. Consensus morphotypes are highlighted in bold and labeled according to Figure 5 and preceded by “CM”, whereas uncertain CM are labeled arbitrarily with letters.

To validate the accuracy and robustness of our models, we used a second cohort of 63 slides from the CUH2 cohort representing a similar spectrum of diagnoses to the MLL cohort but digitized using a different slide scanner (Aperio AT2). In all cases, we evaluated the best performing fold from the previous cross validation. These results show that both our models, glmnet and multi-objective Morphotype analysis, displayed good generalization, with most external validation AUC intervals overlapping with cross-validated AUC estimates (**Figure 6c,d**). We note that including morphotypes found to be statistically unstable in the original discovery step led to a deterioration of validation accuracy in the disease detection task. Further, the single objective Morphotype analysis yielded worse generalization even when limiting to stable morphotypes (**Supplementary Figure S14**), indicating that simultaneously learning multiple tasks unravels more robust morphotypes.

## 4 Discussion

We present an automated protocol for the detection and characterization of thousands of blood cells in PBS linked with machine-learning methods that can use these cellular descriptions to distinguish between clinical conditions and identify novel associations between cytomorphological phenotypes and clinical conditions. Finally, we show that our approach generalizes to other centers and scanners.

Haemorasis, our open-source method to extract and characterize large numbers of WBC and RBC from digitized PBS, demonstrates that blood cell detection and characterization can be automated with no recourse to proprietary equipment or software. We make it publicly available as a Docker container, enabling its straightforward application in a high throughput setting, noting that this can be done with a reasonable amount of compute power (given 8GB of RAM and an NVidia GTX 1080 Ti, typical runtimes for Haemorasis were between 30 minutes and 2 hours per slide). This allowed us to extract millions of RBC and WBC from over 400 PBS, characterizing them in a way that enables the prediction of clinically relevant conditions and identification of relevant cytomorphological features. Using morphometric moments (the mean and variance of morphometric features for each PBS) we show the diagnostic importance of the variance of cellular shape for various conditions. This observation bears similarity to previous reports that associated red cell distribution width with increased risk of AML transformation ^52,55^. It is also worth considering that quantifying morphological variation, especially of subtle features, is likely to be challenging to achieve by visual assessment as it requires an absolute quantification and evaluation of many cells.

We then developed a method that classifies individual cells into disease-associated morphotypes and applied it to over half a million WBC and millions of RBC. This uncovered that MDS cases with a larger prevalence of hypolobulated neutrophils are more likely to harbor *SF3B1* mutations. Also, whilst *SF3B1*-mutant MDS displays characteristic features in the bone marrow, little is known about the differences between *SF3B1*-mutant and *SF3B1*-wildtype MDS regarding RBC morphology in the blood. We also found that RBC size is elevated in *SF3B1*-mutant MDS, as previously suggested for the differential of MCV in *SF3B1*-mutant and *SF3B1-*wildtype MDS ^56^. Additionally, neutrophil hyperlobulation was robustly detectable not only in megaloblastic anemia but also in iron-deficiency ^57,58^ demonstrating that the ability to computationally analyze large numbers of cells can detect this feature even when it is subtle and would otherwise require enumeration of large numbers of neutrophils and their lobe count by experts ^58^. We also observed that neutrophils are larger in megaloblastic anemia, highlighting a common mechanism behind the enlargement of both RBCs and neutrophils in this condition. By analyzing the distribution of different morphometric features, we discovered morphotypes that reflect changes in morphological heterogeneity associated with different conditions. Together, these results highlight how computational cytomorphology can be used to discover RBC and WBC morphotypes that explain observed disease-associated morphological heterogeneity.

Finally, we show that morphotypes are enriched with known cell types and that our approach validates externally, generalizing to PBS from other centers obtained using different slide scanners. However, further studies would be required to validate the described morphotypes in a clinical setting — the expert analysis of PBS considering the morphotypes here discovered would be the ideal validation, and more diverse training and validation cohorts could further confirm the generalization capabilities of automated cytomorphology, which can still be affected by preparation- and scanner-specific artifacts and noise ^59,60^. Efforts like the National MDS Natural History Study, to be concluded in 2025, seek to constitute the first multicentre cohort and tissue-bank for MDS ^61^ and could be used as a more comprehensive assessment of the generalization of computational cytomorphology in the context of MDS. Applying morphotype analyses to larger and more diverse cohorts, both in terms of their clinical preparation, but also comprising a greater variety of conditions is also likely to discover further morphotypes, which in this analysis only account for 60% of WBC and 90% of RBC, and to improve the definition of existing ones.

Our study reveals the potential utility of computational cytomorphology of PBS in the detection of hematological diseases. Additionally, it demonstrates an approach through which researchers can use existing cohorts of annotated PBS to identify novel morphotypes associated with specific clinical conditions and highlights how this can be made generalizable by considering consistent and stable morphotypes. Finally, our study provides proof-of-principle that computational cytomorphology can augment the ability of automated blood cell analyses to identify abnormalities suggestive of hematological disease, at high throughput and minimal additional cost. This may be helpful to identify patients in need of further and usually more invasive and expensive testing, such as bone marrow aspirates or genome sequencing. Recent applications of computational cytomorphology on bone marrow smears have demonstrated its ability to automatically identify different leukocytes ^30,62^ and assist diagnostic predictions ^27–29^ in specialized haemato-oncology. By demonstrating that this can now be extended to blood smears/slides, our work reveals the potential for the large-scale incorporation of automated cytomorphology into routine diagnostic workflows.

## Supporting information

Supplementary Figures, Methods and Results and Supplementary Tables S3 and S4

## Data Availability

https://www.doi.org/10.6084/m9.figshare.19371008

https://www.doi.org/10.6084/m9.figshare.19369391

https://www.doi.org/10.6084/m9.figshare.19153760

https://www.doi.org/10.6084/m9.figshare.19164209

## Authorship

Contribution: M.G., G.S.V. and J.G.A. conceived the project and wrote the manuscript. J.G.A. developed and implemented the project. J.G.A., E.G. and J.C. scanned peripheral blood slides. T.H. provided peripheral blood slides and assisted with scanning and retrieval of blood count data for the Munich Leukemia Laboratory Set. W.G.D. retrieved blood count data. M.B. and E.G. annotated white blood cells.

## Acknowledgements

J.G.A. is supported by the NIHR Cambridge BRC and their opinions are not necessarily those of the NHS, the NIHR or the Department of Health and Social Care. G.S.V. is funded by a Cancer Research UK Senior Cancer Fellowship (C22324/A23015) and work in his lab is also funded by the European Research Council, Kay Kendall Leukaemia Fund, Blood Cancer UK and the Wellcome Trust. The authors would like to acknowledge Drs. Martin Bessner, James Russell and Duncan Brian for their efforts in annotating blood cells.

## Notes

### Competing Interest Statement

G.S.V. is a consultant for Astrazeneca and STRM.BIO. The other authors declare no competing interests.

### Funding Statement

GSV is supported by a Cancer Research UK Senior Cancer Fellowship (C22324/A23015) and work in his lab is also funded by the European Research Council, Kay Kendall Leukaemia Fund, Blood Cancer UK, and the Wellcome Trust. JGA receives funding from EMBL.

### Author Declarations

All Munich Leukemia Laboratory data provided for this investigation were reviewed and approved by Munich Leukemia Laboratory's internal institutional review board and follow the European Union's General Data Protection Regulation (GDPR). This research was conducted in line with the European Molecular Biology Laboratory's internal policy 53 (Internal Policy regarding the Use of Human Biological Material).

