## Supplementary Figures, Methods and Results and Supplementary Tables S3 and S4 for "Computational analysis of peripheral blood smears detects disease-associated cytomorphologies"

### Supplementary Information

#### Supplementary figures

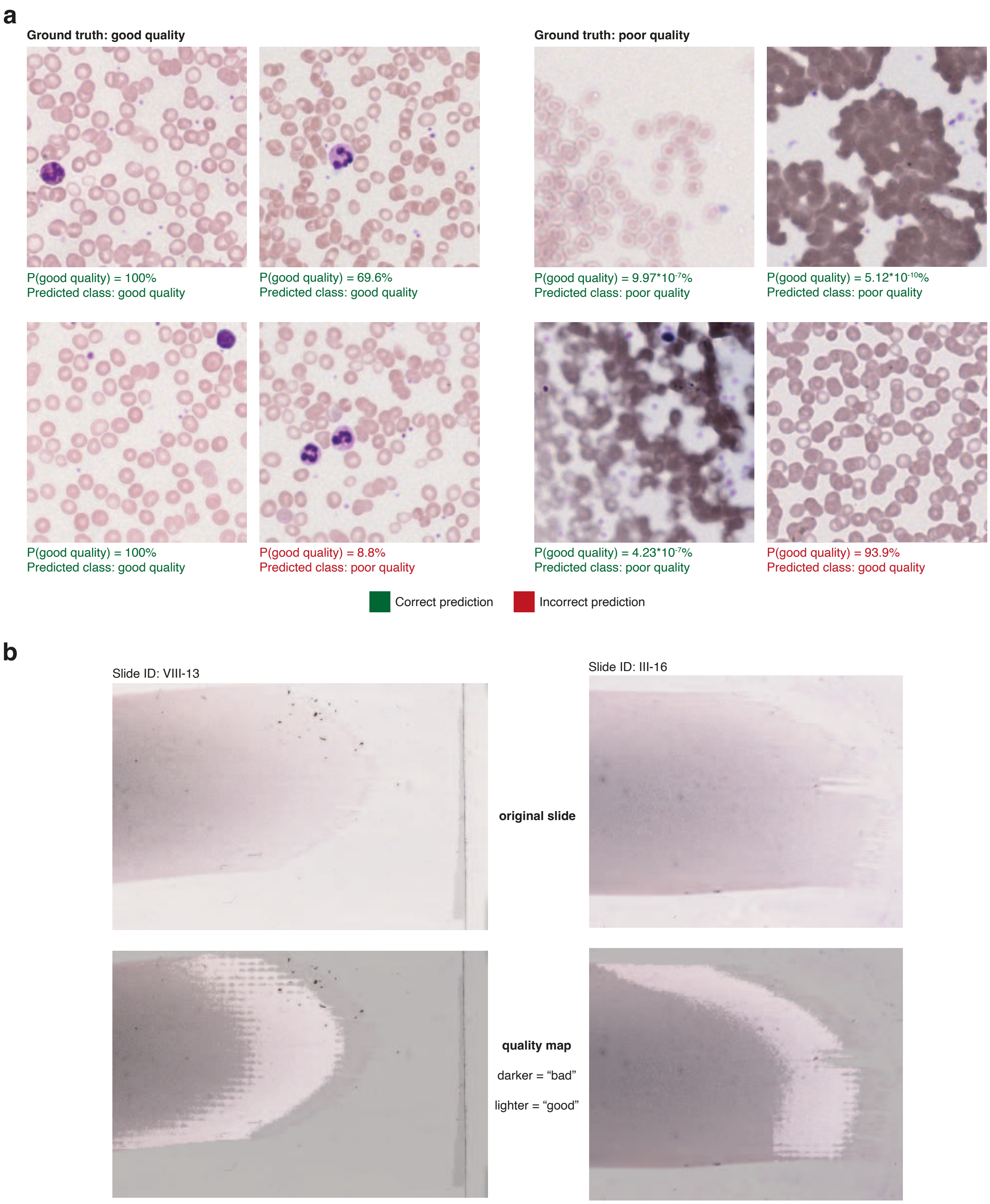


Supplementary Figure S1 - **Quality control examples. a** — Good and poor quality tiles and their respective predictions and prediction probability according to a neural network trained to predict tile quality. **b** — Quality maps (regions of the slide classified as being of good quality) for two PBS.

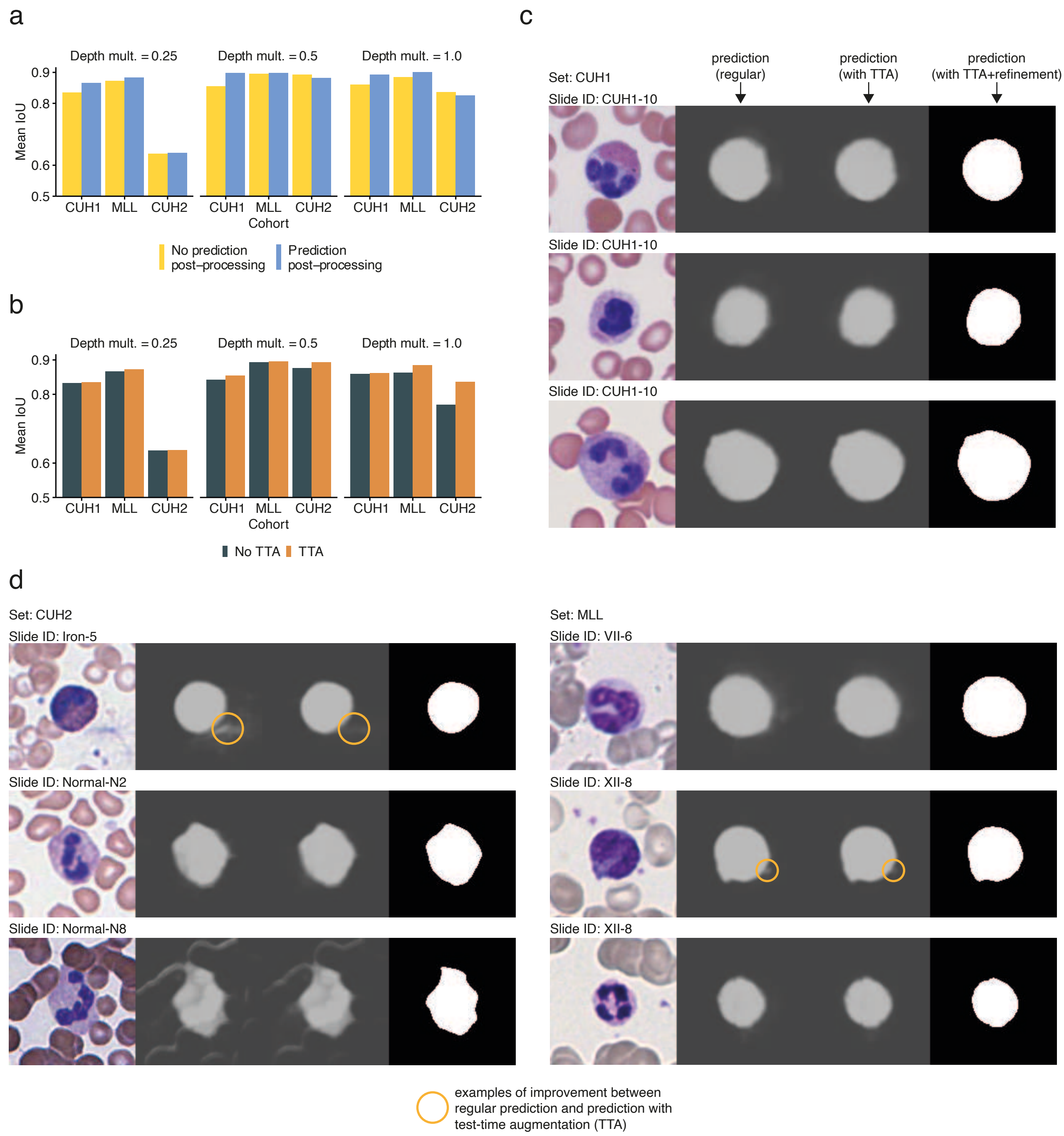


Supplementary Figure S2 - **U-Net WBC segmentation performance. a** — Neural network performance with and without prediction post-processing stratified by network depth and dataset. **b** — Neural network performance with and without test-time augmentation (TTA) stratified by network depth and dataset. **c,d** — WBC segmentation examples. For both **a** and **b**, the training dataset is always CUH1, whereas the other dataset (the MLL dataset and CUH2) are validation dataset.

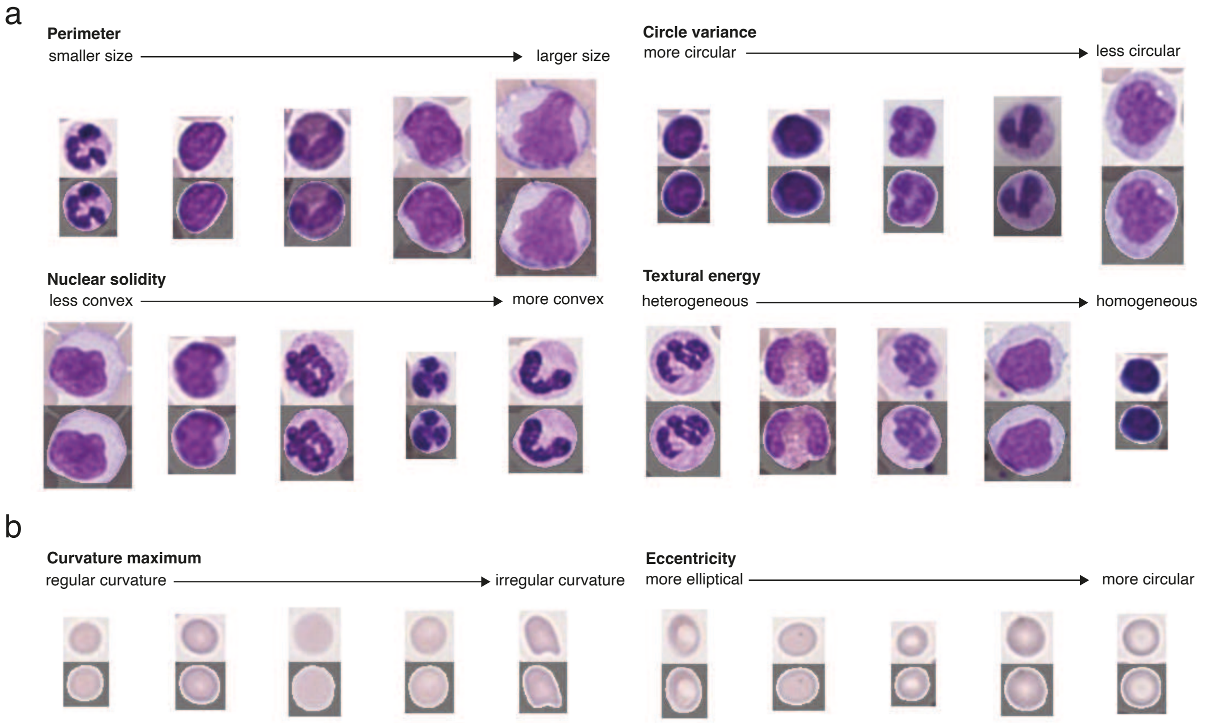


Supplementary Figure S3 - **Examples of cellular characterization using the protocol delineated in this work. a** — WBC examples. **b** — RBC examples.

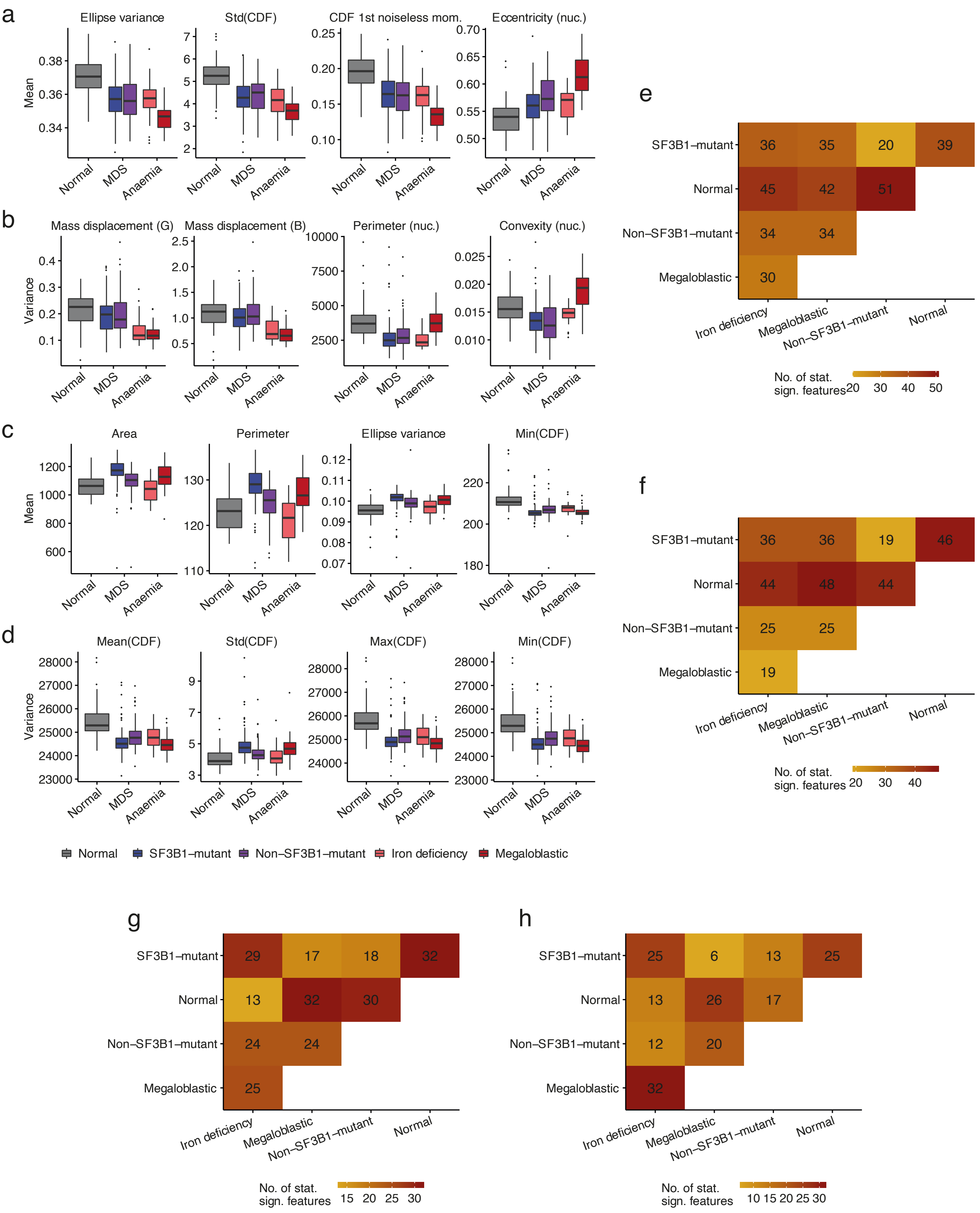


Supplementary Figure S4 - **Individual aspects of morphometric feature distribution carry discriminatory power to predict different conditions. a-d** — Distribution of 4 highly discriminatory feature means in WBC, feature variances in WBC, feature means in RBC and feature variances in RBC, respectively. Highly discriminating features were detected as having the highest Kruskal-Wallis statistic out of all features. **e-h** — Number of highly discriminatory WBC feature means, WBC feature variances, RBC feature means and RBC feature variances, respectively, for all condition combinations (according to significant comparisons in a Dunn-Bonferroni test on features which were significant for a Kruskal-Wallis test).

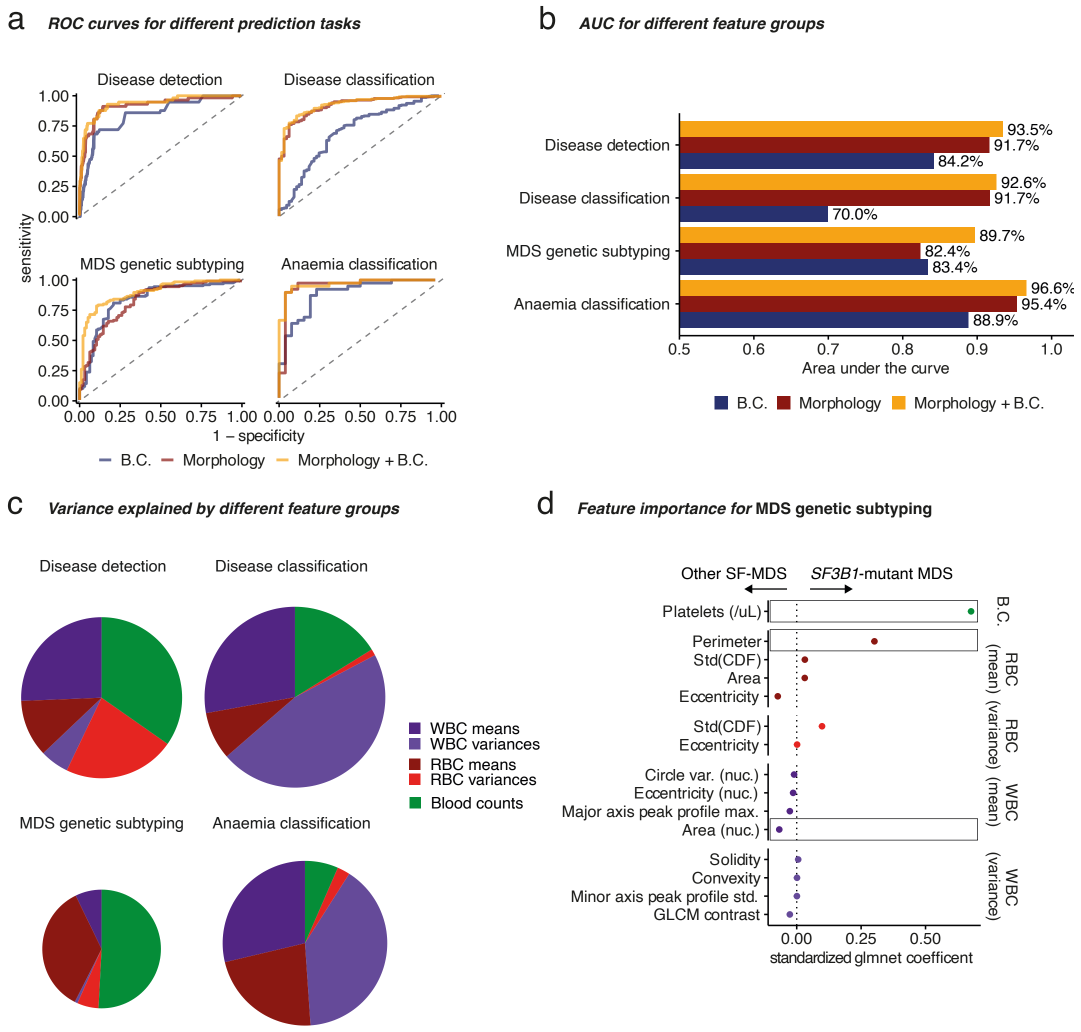


Supplementary Figure S5 - **Automated morphology improves condition prediction.** **a** — Cross-validated receiver operating characteristic (ROC) curves for the four considered tasks. **b** — Cross-validated area under the ROC (AUC) curve. **c** — Feature group contribution to different tasks from 4 models trained on both morphology and blood counts (B.C.). In **b** and **c**, lines and bars are coloured according to the used datasets for each task. In **c**, circles are scaled according to the total explained variance and coloured according to the feature group. **d** — Individual feature importance for MDS genetic subtyping using glmnet.

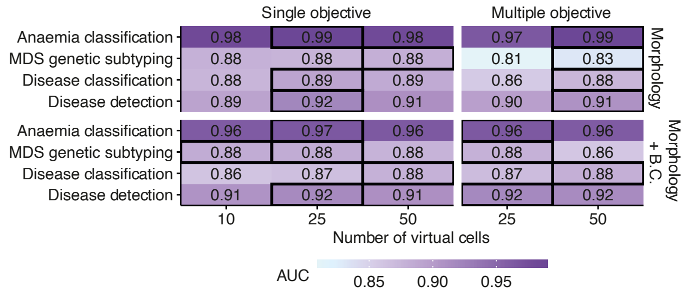


Supplementary Figure S6 - **Performance (AUC) of the multiple and single objective Morphotype analysis model according to the assumed number of computational morphotypes and for different tasks.**

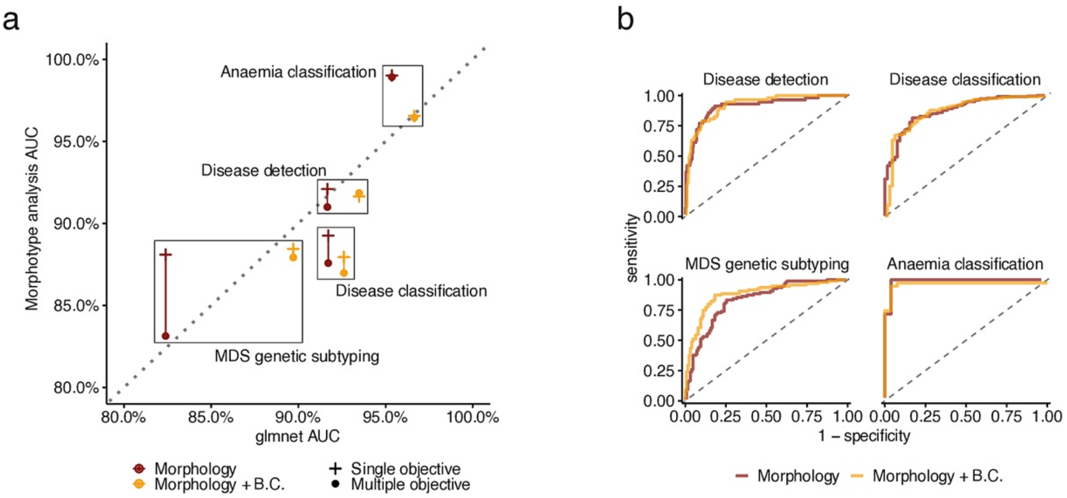


Supplementary Figure S7 - **Predictive performance of morphotype analysis and comparison with glmnet. a** - Comparison of the cross-validated AUC for glmnet and single and multiple objective Morphotype analysis. **b** - Receiver-operating characteristic curves for the predictive performance of Morphotype analysis across different tasks.

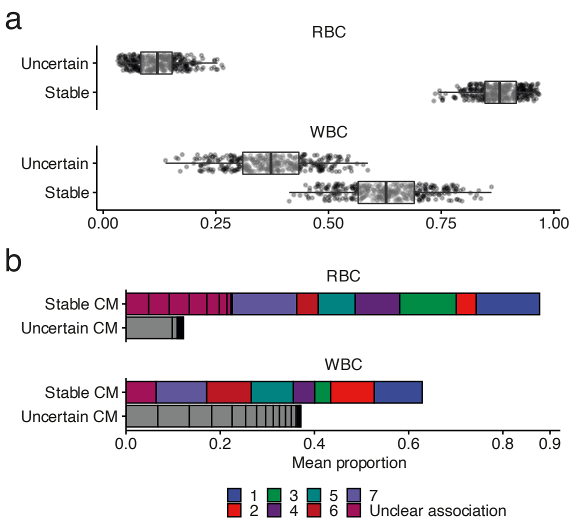


Supplementary Figure S8 - **Morphotype stability in WBC in RBC. a** - Stable and uncertain RCMs (top) and WCMs (bottom) per slide. **b** - Mean proportion of stable RCMs (top) and WCMs (bottom) stratified by CM identity. Colours correspond to different RCMs/WCMs and the bar fractions corresponding to CMs with unclear associations/uncertain CMs are divided by black lines according to the mean prevalence of their constituting CMs.

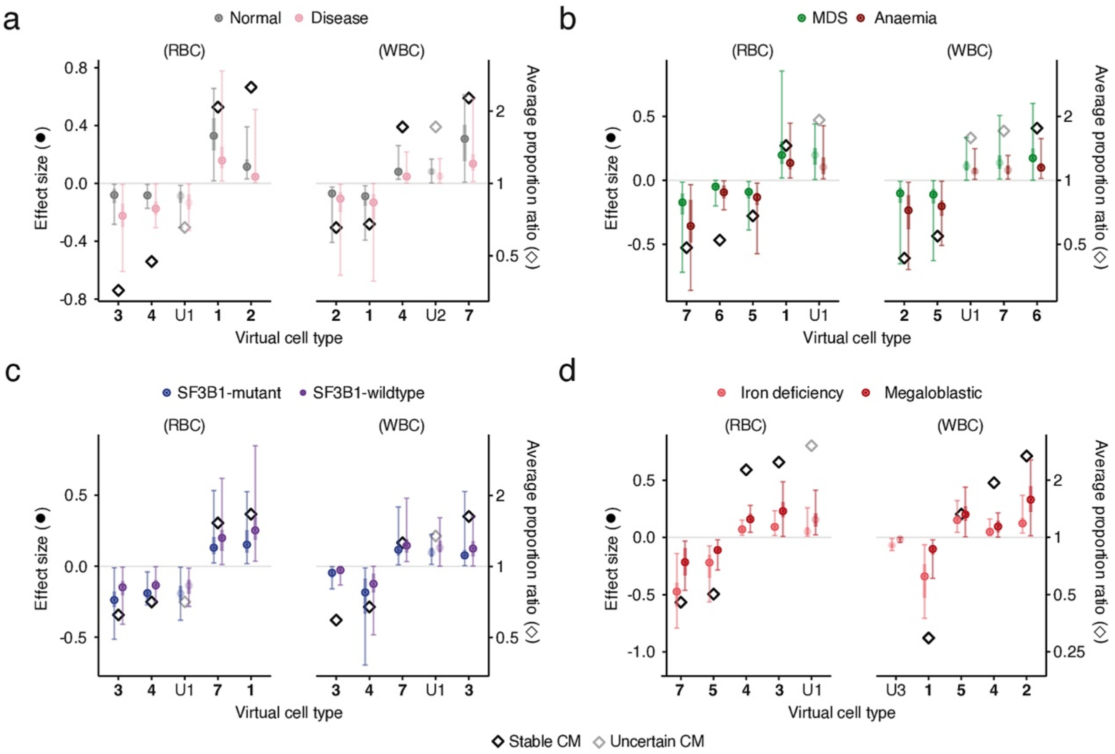


Supplementary Figure S9 - **Effect sizes for the most discriminative computational morphotype (CM) proportions for the multiple objective Morphotype analysis predicting different tasks. a** — Disease detection (normal vs. disease). Effect sizes above zero are biassed towards the “normal” classification. **b** — Disease classification (MDS vs. anemia). Effect sizes above zero are biassed towards the “MDS” classification. **c** — *SF3B1*-mutant detection (*SF3B1*-mutant MDS vs. *SF3B1*-wildtype MDS). Effect sizes above zero are biassed towards the *SF3B1*-wildtype MDS classification. **d** — anemia classification (iron deficiency vs. megaloblastic). Effect sizes above zero are biassed towards the “megaloblastic” classification. For a-d, the effect size was calculated as the product of the proportion of each CM and its coefficient in Morphotype analysis. Each point (circle) represents the median effect size and the error bars represent the 90% confidence interval for the effect sizes and corresponds to the left y-axis. Each diamond represents the proportion ratio for each condition and corresponds to the right y-axis.

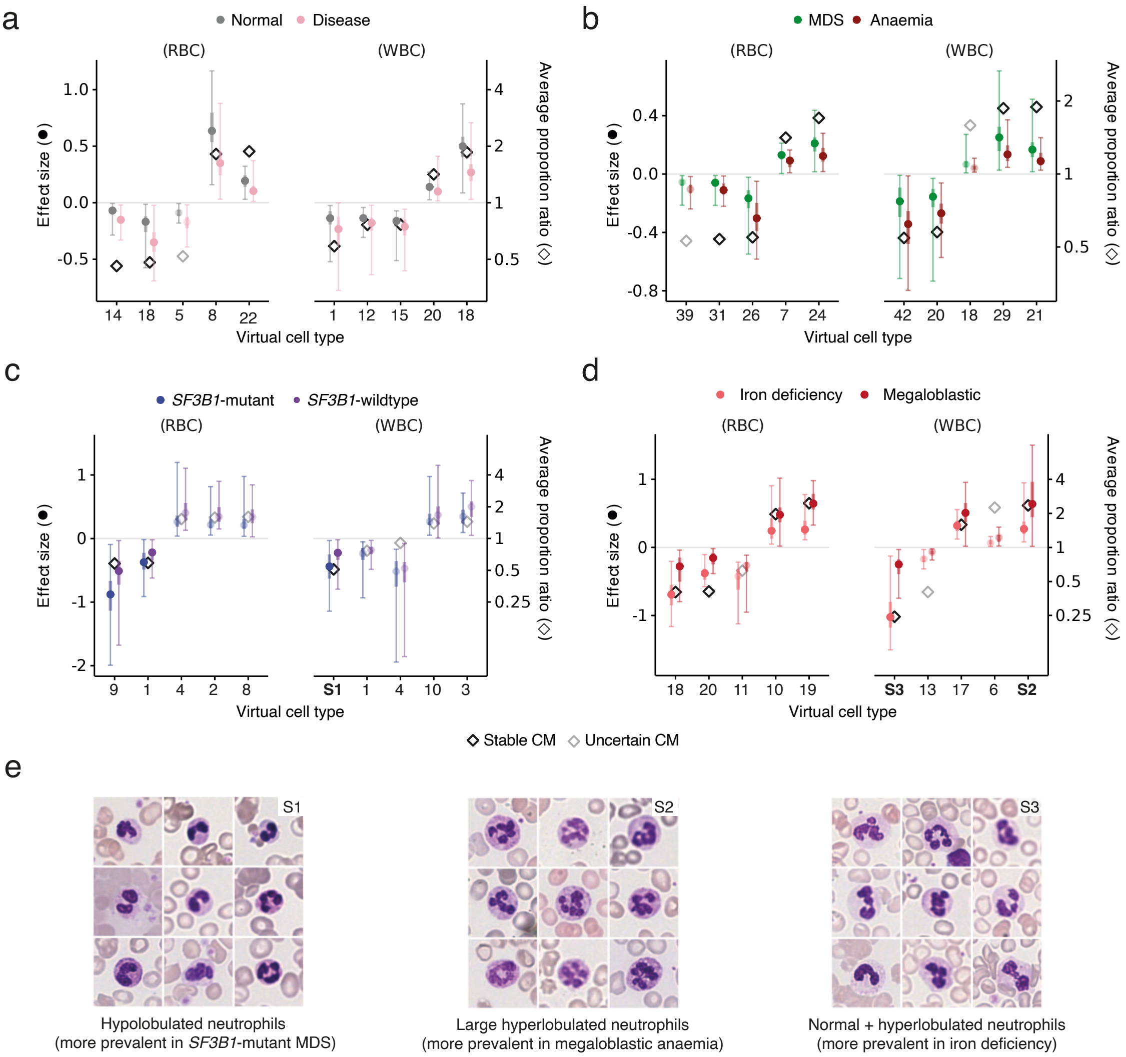


Supplementary Figure S10 - **Effect sizes for the most discriminative computational morphotype (CM) proportions for the single objective Morphotype analysis models. a** — Disease detection (normal vs. disease). Effect sizes above zero are biassed towards the "normal" classification. **b** — Disease classification (MDS vs. anemia). Effect sizes above zero are biassed towards the "MDS" classification. **c** — *SF3B1*-mutant detection (*SF3B1*mutant MDS vs. *SF3B1*-wildtype MDS). Effect sizes above zero are biassed towards the "SF3B1-wildtype MDS" classification. **d** — anemia classification (iron deficiency vs. megaloblastic). Effect sizes above zero are biassed towards the "megaloblastic" classification. For **a-d**, the effect size was calculated as the product of the proportion of each CM and its coefficient in Morphotype analysis. Each point (circle) represents the median effect size and the error bars represent the 90% confidence interval for the effect sizes and corresponds to the left y-axis. Each diamond represents the proportion ratio for each condition and corresponds to the right y-axis. **e** — Examples of CMs validating some conclusions from SO Morphotype analysis.

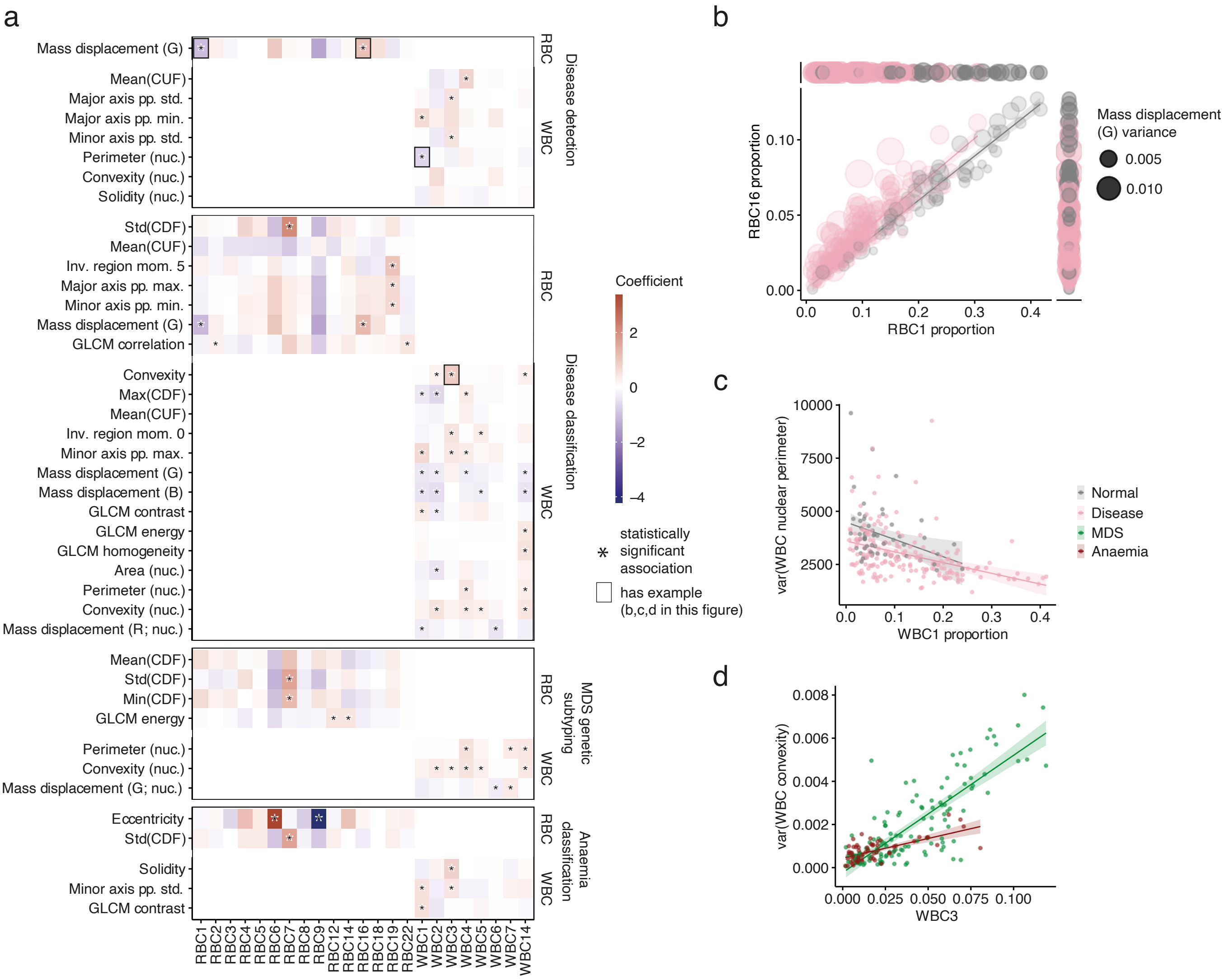


Supplementary Figure S11 - **Computational morphotype proportions capture broad morphometric trends. a** — Partial associations between CM proportions (x-axis) and the variances of different morphometric features (2nd morphometric moment; y-axis). Partial associations (coefficients) between consensual CM and the variance of specific features were calculated according to a linear model that $var_f = \sum_i^n (cCM_i \times b_i) + a$, where $var_f$ is the variance of feature $f$, $n$ is the total number of consensual CM, $cCM_i$ is the proportion of consensual CM $i$, $b_i$ is the coefficient for proportion $cCM_i$ and $a$ is an intercept term. The statistical significance of coefficient estimates was assessed using a two-sided t-test. **b** — Relationship between RCM types 16 and 7 (the latter is A in Figure 5c and Figure 6b) and the mass displacement variance (size of points) stratified by slide classification according to the disease detection model (normal vs. disease). **c** — Relationship between WCM 14 (C in Figure 5b and Figure 6a) and the variance of the WBC nuclear perimeter stratified by slide classification according to the disease detection model (normal vs. disease). **d** — Relationship between WCM 11 (G in Figure 5b and Figure 6a) and the variance of the WBC convexity stratified by slide classification according to the disease classification model (MDS vs. anemia). All reported CMs are stable. CM1-7 in both WCM and RCM are consistent with those reported in previous figures. Other CMs have been arbitrarily numbered.

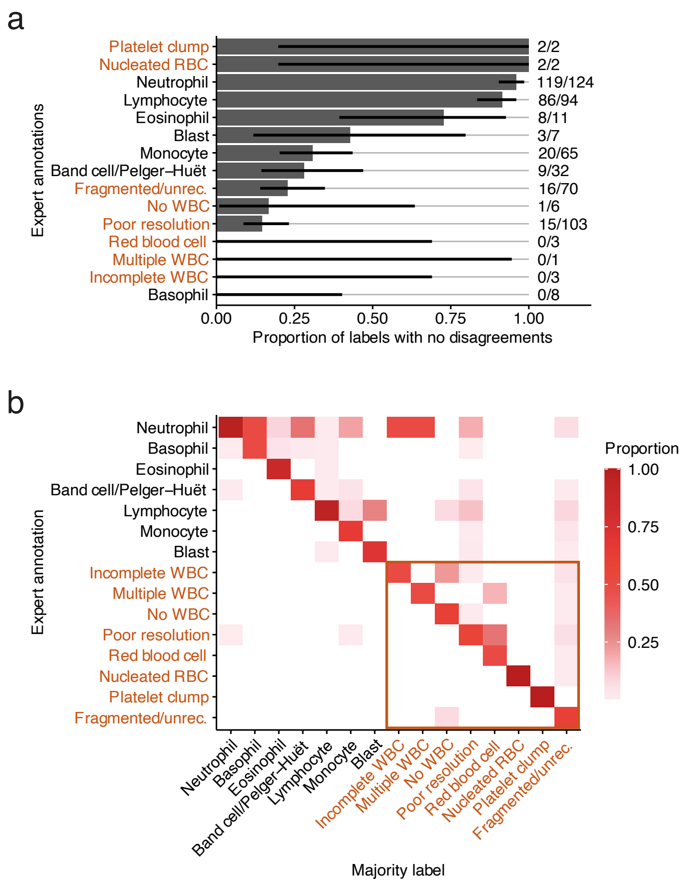


Supplementary Figure 12 - **Concordance between experts in cell annotation. a** — Proportion of expert-labelled WBC with no disagreements when cells were labelled by at least two experts. **b** — Confusion matrix comparing the majority label with the fraction of experts annotating each cell as the majority label.

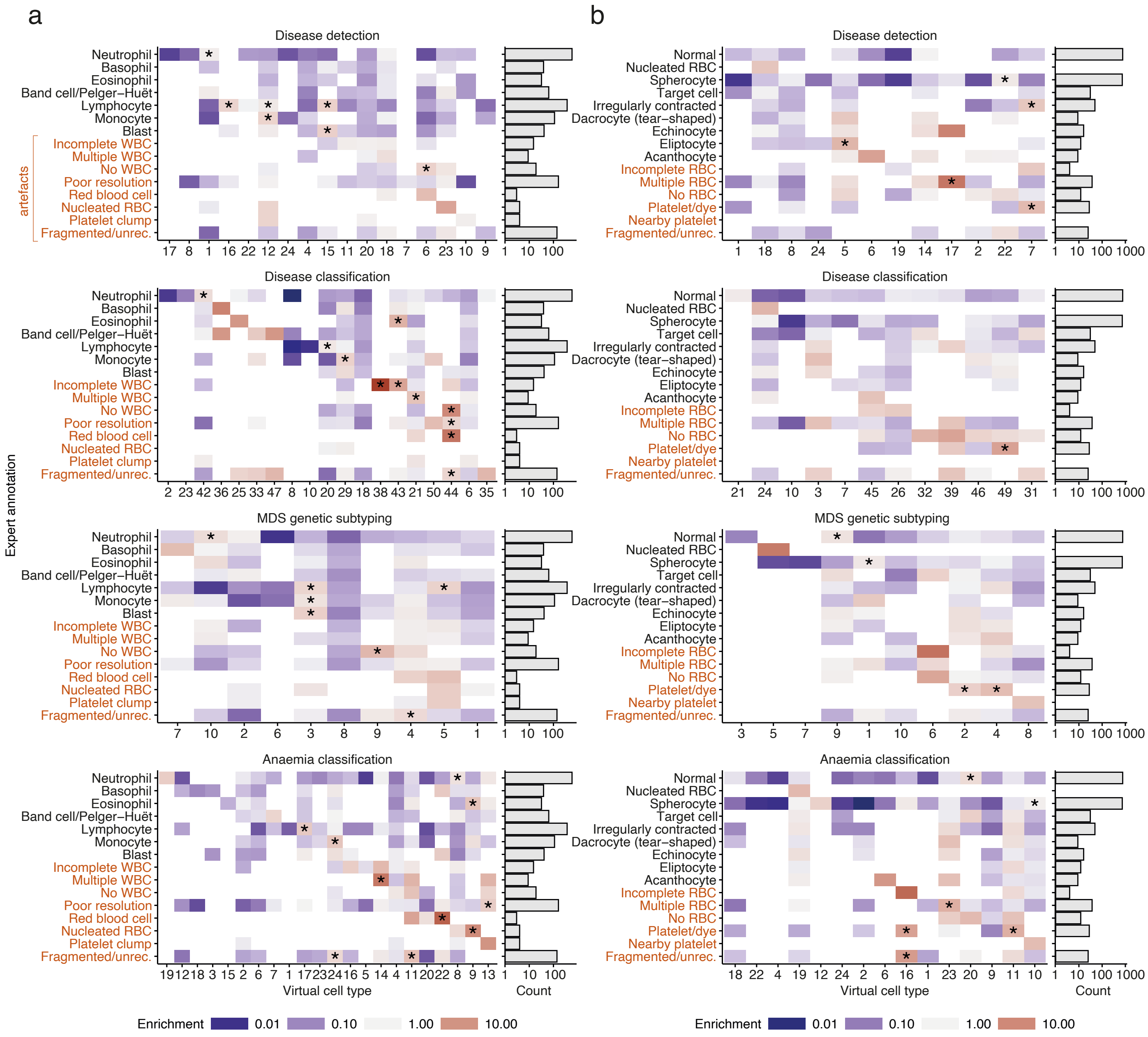


Supplementary Figure 13 - **Computational morphotypes capture expert-annotated cell types with no previous annotation in single objective MILCoMorI models. a** — Correspondence between WCM types and expert-annotated WBC types (left) and number of annotated WBC types. **b** — Correspondence between RCM types and expert-annotated RBC types (left) and number of annotated RBC types.

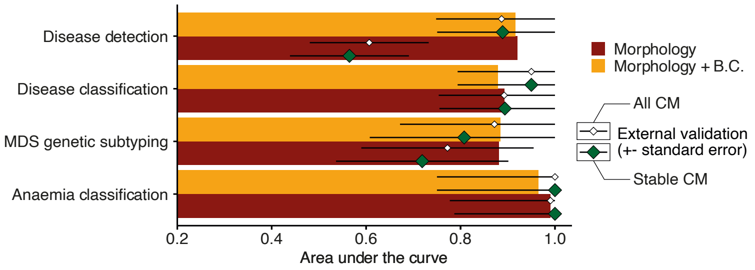


Supplementary Figure 14 **- External validation for single objective Morphotype analysis models.**

### Supplementary Methods

#### PBS tiling for prediction

Analysing the entire PBS at once using either RBC or WBC detection protocols is infeasible due to memory constraints — if each pixel is represented as a set of 3 8-bit integers (as is usually the case with images) a PBS from the MLL dataset represents between 11GB and 75GB of required memory; the first layer of the U-Net, with 32 bit output channels, could require 797GB of memory, an unfeasibly high requirement for most computers. For this reason, we consider individual slide tiles which, in our case, are $512 \times 512$ pixel for the quality control network and $640 \times 640$ pixel ($512 + 64 \times 2$) for RBC and WBC detection. To avoid predicting, at any given point, cells which may be incomplete in a given tile, we exclude all predictions with pixels close to the tile border and to avoid missing these cells use a sliding window with stride 128. Considering that this may lead to detecting the same cell twice as the same object, we exclude a prediction if a cell has already been predicted within 8 pixels. Through this, we estimate the memory requirements for Haemorasis — video or regular RAM — to be 8GB, manageable by most computers and low-to-mid end GPUs.

#### Machine-learning metrics

To evaluate the performance of the quality control, RBC filtering, WBC segmentation and condition prediction, we first define the possible outcomes of a prediction in terms of their correctness and class:

- True positive (TP) — a prediction is correct and belongs to the positive class
- True negative (TN) — a prediction is correct and belongs to the negative class
- False positive (FP) — a prediction is incorrect and belongs to the positive class
- False negative (FN) — a prediction is incorrect and belongs to the negative class

We can then define the metrics utilized over this work:

- Precision — the ratio of TP to predicted positives (TP + FP): $\frac{TP}{TP+FP}$
- Recall (or sensitivity) — the ratio of TP to total positives (TP + FN): $\frac{TP}{TP+FN}$
- Specificity — the ratio of TN to total negatives (TN + FP): $\frac{TN}{TN+FP}$
- Accuracy — ratio of correct predictions (TP + TN) to all predictions (TP + TN + FP + FN): $\frac{TP+TN}{TP+TN+FP+FN}$
- Intersection over union (IoU) or Jaccard score — the ratio of TP to the union of true positives and predicted positives (TP + FP + FN): $\frac{TP+TN}{TP+TN+FP+FN}$. This can also be stated as the ratio between the intersection of true positives and predicted positives and the union of true positives and predicted positives.
- $F_1$ — the harmonic mean of the precision and recall: $\frac{2}{\frac{1}{recall}+\frac{1}{precision}} = \frac{precision \times recall}{precision+recall}$. This metric has the advantage of, unlike accuracy, provide a more realistic assessment of predictive performance that is less affected by cases of imbalanced data

We also use the area under the receiver operating characteristic (ROC) curve (AUC). This curve is calculated using the class probabilities $p$ produced by a given predictive model. Using a sequence of thresholds between the minimum and the maximum of $p$, different values for the sensitivity and specificity are calculated, and a trapezoid curve is constructed where each point corresponds to a threshold value and its coordinates are $\{1 - specificity, sensitivity\}$. The area under this curve (the ROC curve — AUC) is thus a balanced measure of how well a model performs that is insensitive to model calibration.

#### Quality control network

This step intends to exclude tiles which are either blurred, have very high cellular density or very low cellular density. While there are relatively simple ways to quantify "blurriness" through a proxy (calculating the variance of the Laplacian, for instance), accurately identifying scenarios of very high or very low cellular density is not as trivial since it requires contextual information (which kinds of cells are visible in the image). Recent work has shown that deep-learning can accurately quantify the blurriness of an image in microscopy settings, outperforming other *ad hoc* metrics devised to quantify blurriness [63].

To do this, we trained an ImageNet-pretrained DenseNet121 model for 25 epochs with a batch size of 32 and the Adam optimizer with an initial learning rate of 0.00005 that decayed by 90% every time the training loss stopped decreasing. During training, each image had a 60% probability of having its brightness, saturation, hue and contrast randomly altered (by 15%, 10%, 10% and 10%, respectively) or of having random JPEG compression artefacts introduced. This makes the training more robust to alteration of PBS preparation and variability in image digitalization.

#### Red blood cell segmentation

The detection of RBC is performed using a routine described in Algorithm 1. For clarity, $CannyEdgeDetector$ represents the Canny edge detection algorithm [33], $getContours$ is a standard routine to detect object contours from a binary image, $drawContours$ is a routine to draw contours on the image and fill them, $getArea$ is a function that calculates the area of a contour or ellipse, $averageColour$ is a function that selects the pixels in an image belonging to a contour and calculates their average value, $fitEllipse$ is a function that fits an ellipse to a contour, $getMajorAndMinorAxis$ is a function that calculates the lengths of the major and minor axes of an ellipse and $isolateObject$ is a function that returns the region of the image containing the RBC and its respective mask.

#### Data augmentation and prediction-time enhancements to U-Net predictions

To train the U-Net models we used data augmentation to achieve the best possible results — using the original $2096 \times 2096$ pixel images, $512 \times 512$ pixel tiles are randomly extracted after rotating the image at a random angle between 0 and 89 degrees. Next, each image is distorted using an elastic transform as implemented in albumentations [64], following random image flips and/or rotations and perturbations to the brightness, saturation, hue and contrast of the image. Finally, we randomly add salt and pepper and Gaussian noise to each image, which can also be slightly blurred (using a standard Gaussian blur). The probabilities and parameters for these augmentations are presented in **Supplementary Table S3**.

To ensure the best possible performance using our U-Net model, we used test-time augmentation (TTA) and prediction post-processing. The former is characterized as rotating and flipping the input image in order to derive a consensus prediction from the several outputs, whereas the latter was a custom process. Particularly, for TTA, we rotate the image by 90, 180 and 270 degrees (4 inputs) and then rotate the outputs to the original image orientation, calculating the average pixel probability across the 4 outputs [37]. Prediction post-processing is done by removing objects detected as WBC whose size lies outside the expected distribution for WBC sizes, and filling small convex hull defects.

**Data:** $image$
**Result:** Set of masked RBC images and their contours $S$
$S \leftarrow \{\}$;
$edgeImage \leftarrow CannyEdgeDetector(image)$;

```
contours ← getContours(edgeImage);
contourImage ← drawContours(contours);
contourImage ← contourImage − edgeImage;
contours ← getContours(contourImage);
for contour in contours do
  area ← getArea(contour);
  if area > 300px and area < 2000px then
    averageColour ← getAverageColour(image; contour);
    if averageColour > 170 and averageColour < 220 then
      ellipse ← fitEllipse(contour);
      majorAxis, minorAxis ← getMajorAndMinorAxis(ellipse);
      areaEllipse ← getArea(ellipse);
      if areaEllipse < 1500px and areaEllipse > 300px and majorAxis/minorAxis < 1.5 then
        isolatedCellImage; isolatedCellMask ← isolateObject(image; contour);
        append {isolatedCellImage, isolatedCellMask, contour} to S;
      end
    end
  end
end
```

**Algorithm 1:** Red blood cell segmentation and detection algorithm.

#### Blood counts and morphometric moment preprocessing and feature importances in glmnet

Before training the elastic net models, we standardize variables and impute missing values in the blood counts using the median value of each fold (a total of 139 blood counts were inferred for 49 individuals). To assess feature importance in glmnet, we use the absolute value for the standardized coefficients. To calculate the feature importance for the 5 relevant groups of features — for the means and variances of WBC (2) and RBC (2) and the blood counts (1) — we first calculate the effect of each group of variables by summing the effects of the standardized variables composing it for each individual. Then, we calculate the total explained variance by each group of features by calculating the sum of the rows in the covariance matrix between all 5 groups of features and dividing it by the total explained variance (sum of all elements of the covariance matrix).

#### Morphotype analysis — model specification and optimization

**Model specification.** A given PBS is characterized as a set of $n$ cells $O \in R^{n\times f}$, where $f$ is the number of morphological features characterizing each cell. We want to assign each of the $n$ cells to one of $v$ CM, such that $V = softmax(O \times \theta_V, axis = 1$, where $V \in R^{n\times f}$ is the membership (between 0 and 1) of all cells to each CM and $\theta \in R^{f\times v}$ are the parameters of the softmax regression assigning each cell to one of $v$ CM. The computational morphotype (CM) composition of the slide $C \in \Delta^v$, where $\Delta^v$ is the simplex of order $v$, is then calculated as $C = mean(V, axis = 0)$. $C$ is then used to calculate the probability that a PBS belongs to an individual with a given disease $d$ as $P(d|C) = sigmoid(\theta_C \times C)$, where $\theta_C$ parametrizes the weights of each CM for the classification probability $P(d|C)$.

We characterize the RBC $R \in R^{r\times f}$ and WBC $W \in R^{w\times f}$ in a slide $S$ as separate entities, where $r$ and $w$ are the number of RBC and WBC, respectively, and $f$ and $g$ are the number of features characterizing each RBC and WBC, respectively. Then, the RBC and WBC CM (RCM and WCM, respectively) compositions of each slide are calculated separately (with $\theta_{R,RBC} \in$, $R^{f\times v}$ and $\theta_{W,WBC}, \in R^{g\times v}$, respectively) and use them to calculate $P(d|C_{RBC}, C_{WBC}) = sigmoid(\theta_{C,RBC} \times C_{RBC} + \theta_{C,WBC} \times C_{WBC})$. This formulation also enables the use of additional information, particularly blood counts ($counts$, used earlier in this chapter) such that $P(d|C_{RBC}, C_{WBC}, counts) = sigmoid(\theta_{C,RBC} \times C_{RBC} + \theta_{C,WBC} \times C_{WBC} + \theta_{counts} \times counts)$.

Model optimization. We are interested in the concurrent optimization of $\theta_{V,RBC}$, $\theta_{V,WBC}$, $\theta_{C,RBC}$, $\theta_{C,WBC}$ and $\theta_{counts}$ such that Morphotype analysis models classify cells into condition-specific CM while learning how to predict conditions from specific PBS. For this reason, we used stochastic gradient descent to optimize Morphotype analysis models, assuming that there are an equal number of WCM and RCM. We use a cross-entropy loss and $L_2$ regularization where λ = 0.2.

We also train multiple objective models which optimize all four tasks using the same set of CM. For this, in each training step and for each of the four tasks, a set of relevant PBS is sampled and the cross-entropy is calculated. All four loss values are summed after multiplying them by a vector sampled from a Dirichlet distribution with concentration [1,1,1,1] as proposed in [65] to approximate Pareto optimality (i.e. no improvement can be done to one task without making a different task worse [66]) and backpropagate this value.

To select the best hyperparameter values (number of CM) in each setting we used the cross-validated AUROC. To get these cross-validated scores, each model is trained over 5 folds and the average value and standard error of each metric is calculated. In each fold, a model is optimized for a maximum 15,000 steps with a learning rate of 0.01 and a weight decay of 0.25. We used weighted cross-entropy with the Adam optimizer [32]. The weight for the positive class is calculated as $1 - \frac{no.\ of\ elements\ in\ the\ positive\ class}{no.\ of\ total\ elements}$. Additionally, the learning rate is decreased every time the training loss stagnates for at least 100 steps by a factor of 0.5. If the learning rate reaches a value of 0.000001 (smaller than the initial learning rate by 4 orders of magnitude) training is stopped. Each training iteration samples $500$ WBC and $500$ RBC. This may lead to a situation where bags of cells with a smaller amount of information (fewer cells) contribute equally to those with a large amount of cells. To minimize this, each instance is weighted with $w_{avail}$, calculated from the number of available RBC $m$ and WBC $n$ such that $w_{avail} = 1 + \frac{minimum(500,m)+minimum(500,n)}{1000}$. $w_{avail}$ will be closer to 1 when fewer cells are available and, if more than 500 WBC and 500 RBC are available, it will be 2.

We trained models for each task using different numbers of CM — $[25,50]$ for MO Morphotype analysis and $[10,25,50]$ SO Morphotype analysis — assuming that both WBC and RBC can be clustered into the same number of CMs, and whether blood counts (WBC counts (cells/$\mu$m), haemoglobin concentration (g/dL) and platelet counts (platelets/$\mu$m)) improved the classification performance of Morphotype analysis. All variables were standardized (mean = 0 and standard deviation = 1).

**Predictions using stable CMs.** Since we use a single solution (i.e. a single fold) for inference and testing this can lead to a generic problem of determining a latent space (CMs, in our case) — oftentimes, the mapping between input and latent space becomes too specific for the training data on each fold and underperforms on other datasets by detecting CM which correspond to artefacts rather than to clinically-relevant cytomorphologies. For this reason, we use only CM which are stable across the 5 cross-validation folds. To define these CM, we sample 500 WBC and RBC from each PBS with replacement (172,000 WBC and 172,000 RBC in total) and calculate their CM probability (the probability that each cell belongs to a specific CM) for the best performing and other folds ($C_{WBC/RBC,best}$and $C_{WBC/RBC,k}$, respectively, where $best$ corresponds to the best-performing fold and $k \in \{1,2,3,4\}$ corresponds to other folds). We then calculate the correlation $R$ between all CM in the best-performing fold and all CM in other models. This measures how similar a CM in the best performing fold is to those in other folds. Next, we match CM in the best-performing fold to those in other folds by considering, for each CM in each combination of $\mathrm{best}$ and $k$, the CM pair with the highest $R$. Finally, we consider that a cell in $\mathrm{best}$ is stable whenever there are three or more matches where $R > R_{thresh}$ (in our case, $R_{thresh} = 0.5$). In other words, a CM is stable whenever it is sufficiently correlated with any other CM across most folds trained with different subsets of the same data.

Inference using stable CMs is then performed by setting the proportions of all non-consensual CM to 0 and distributing the proportions across the remaining, consensual CM. While the ideal case scenario would be to retrain the model with the subset of consensual CM, we only assess performance using the AUC, which does not depend on the absolute value of the classification probability, depending rather on the ranking of different classification probabilities.

**Stable CM labelling in this manuscript.** In **Figure 4**, **Figure 5**, **Supplementary Figure S8** and **Supplementary Figure S9** we show only stable CMs and enumerate them accordingly (WBC CM labels and RBC CM labels correspond with each other across these figures). In **Figure 6**, where we show all CMs enriched in an expertlabelled cell type, we label stable CMs as **CMx**, where **x** corresponds to the numbers in the figures previously stated in this paragraph, while other other CMs are labelled using letters to avoid confusion. In **Supplementary Figure S9**, non-stable (uncertain) CMs are labelled as **Ux**.

**Associations between CM proportions and variance.** To calculate how the relative prevalence of different CM is associated with changes in the variance of specific morphometric features, we fit, for important variance features in the glmnet models (non-zero variance features), the linear model $var_f = \sum_i^n \quad (cCM_i \times b_i) + a$, where $var_f$ is the variance of feature $f$, $n$ is the total number of consensual CM, $cCM_i$ is the proportion of consensual CM $i$, $b_i$ is the coefficient for proportion $cCM_i$ and $a$ is an intercept term. To make each term more comparable, we normalize variance and CM proportions independently as described in "Morphometric moment preprocessing and feature importances in glmnet" in the **Supplementary Methods**. To assess whether these coefficients (associations) are statistically significant, we use t-tests and correct for multiple testing using the Bonferroni method considering a threshold of $\alpha = 0.05$ ($\alpha_{corrected} = \frac{0.05}{405\ tests} = 0.000123$). To calculate the feature densities shown in **Figure 5**, we use a subset of 31,119 RBC and 31,884 WBC from the MLL cohort.

#### Setting up Haemorasis

**As a Docker container.** This is the recommended route as it requires only a Docker installation [67] (also easily transferrable to Singularity) and prevents possible conflicts. The commands to be followed are standard for prebuilt Docker images, i.e. a pre-built image should be pulled from Docker Hub — in this case, `docker pull josegcpa/blood-cell-detection:latest`. This image contains the necessary dependencies and scripts to run Haemorasis.

**From the git repository.** This is not the recommended route as several dependencies have to be installed and errors/incompatibilities are more likely. This route requires the user to have Git, as well as Python 3.6.8 installed. Setting up Haemorasis should be done as follows, assuming the user is operating a UNIX based system and from the command line:

1. Clone the https://github.com/josegcpa/haemorasis repository to the machine where Haemorasis is to be run (`git clone https://github.com/josegcpa/haemorasis`)
2. Install the required packages using pip (`pip install -r requirements-pipeline.txt`)

#### Running Haemorasis

**Simplified explanation of Haemorasis.** Haemorasis is composed of a set of 7 steps, orchestrated using Snakemake:

1. The model checkpoints and parameters are downloaded if they are not available

2. Quality control of the PBS — each $512 \times 512$ pixel tile is quality controlled to filter out tiles with excessive/deficient cellular density and/or poor resolution (output stored as {output.directory}/_quality_control/{slide. id}.h5)
3. Segmentation of WBC and RBC — segmentation coordinates are stored in hdf5 format, with a dataset for each cell (output stored as {output.directory}/_segmented_wbc/{slide.id}.h5 and {output. directory}/_segmented_rbc/{slide.id}.h5, respectively)
4. Morphometric characterization of WBC — morphometric features are stored in hdf5 format, with a dataset for each cell (output stored as {output.directory}/_aggregates_wbc/{slide.id}.h5)
5. Morphometric characterization of RBC — morphometric features are stored in hdf5 format, with a dataset for each cell (output stored as {output.directory}/_aggregates_rbc/{slide.id}.h5)
6. Annotation of RBC in geojson format — these annotations can be loaded into QuPath (output stored as {output.directory}/_annotations_rbc/{slide.id}.h5)
7. Annotation of WBC in geojson format — these annotations can be loaded into QuPath (output stored as {output.directory}/_annotations_wbc/{slide.id}.h5)

Steps 4 and 5 are run in parallel, as well as steps 6 and 7. In the examples above, {slide.id} is the basename of the slide until the first dot (if the slide path is /homes/user/slide_32.0.1.tiff then {slide.id} is slide_32). The output directory ({output.directory}) is the directory which will contain the output from the analysis.

**Pipeline usage — single slide.** To run Haemorasis for a single slide, the run-slide.sh script is used, where two key options are necessary:

- -i — the path to a slide
- -o — the path where the output will be stored

As such, an example of running Haemorasis is, for a slide in /homes/user/slides/slide_a.tiff and the output directory in /homes/user/output is sh run-slide.sh -i /homes/user/slides/slide_a.tiff -o /homes/user/output. sh run-slide.sh -h displays other available options.

**Pipeline usage — folder of slides.** To run Haemorasis for a folder of slides, the run-folder.sh script is used, where three key options are necessary:

- -i — the path to a folder containing slides
- -f — the file extension for the slides (.tiff, .svs, .ndpi and other formats supported by Openslide [68])
- -o — the path where the output will be stored

As such, an example of running Haemorasis is, for a folder of slides in .tiff format in /homes/user/slides and the output directory in /homes/user/output is sh run-folder.sh -i /homes/user/slides -f tiff -o /homes/user/output. sh run-folder.sh -h displays other available options.

**Adaptation to Docker container.** Please note that the examples provided below do not incorporate the Docker command line interface syntax. Additionally, for our use case, where both input and output should be or become locally available, bind mounts are required for the input and output.

As such, to run the Haemorasis command sh run-slide.sh -i /homes/user/slides/slide_a.tiff -o /homes/user/output inside our pulled Docker image josegcpa/blood-cell-

`detection:latest`, one should run `docker run -v /homes/user/slides:/slides -v /homes/user/output:/output josegcpa/blood-cell-detection:latest sh run-slide.sh -i /slides/slide_a.tiff -o /output`. For more information on Docker please consult the Docker documentation in https://docs.docker.com/ and to get started with Docker please refer to the Get started tutorial in https://docs.docker.com/get-started/.

**Visualizing detected cells.** QuPath is a popular software for digital pathology images [50]. Haemorasis has the advantage of producing GeoJSON files with RBC and WBC annotations which can be easily visualized with slides using QuPath in `{output.directory}/_annotations_rbc` and `{output.directory}/_annotations_wbc`, respectively.

### Supplementary Results

#### Analysis of the relevance of individual morphometric features in prediction

To assess whether individual morphometric features (Methods) could be used to discriminate between conditions, we first calculated, for each slide, its morphometric moments — the mean and variance of each morphometric feature obtained for all WBC and RBC — and then assessed whether relevant differences were present between different conditions using a Kruskal-Wallis test. Upon statistical significance after controlling for false-discovery (95%), we determine their utility on specific comparisons using Dunn-Bonferroni tests. We observed that, for WBC, the morphometric moments of 60.0% (36/60 for *SF3B1*mut MDS detection) to 81.7% (49/60 for disease detection) of features are useful to discriminate between different conditions, while for RBC these proportions were between 48.8% (20/41 for disease classification) and 90.2% (37/41 for anemia classification; **Supplementary Figure S4e,h**).

#### Supplementary tables

Table S3: Augmentations used to train U-Net.

| Augmentation | Explanation | Probability | Parameters |
|---|---|---|---|
| **Elastic transform** | A grid is randomly distorted using random gaussian noise leading to slight morphological changes in the cells | 30% | $\sigma = 10$, no affine transformation |
| **Gaussian blur** | The image is blurred randomly using a convolution with side $2 \times size + 1$ | 0.1% | $\mu = 0, \sigma = 0.005, size = 1$ |
| **Random brightness (add.)** | A random value within range is added to all the channels in the image | 75% | $range = [-0.125, 0.125]$ |
| **Random saturation (mult.)** | The saturation (colourfulness of the image) is change by multiplying it by a random value within range | 75% | $range = [0.7, 1.3]$ |
| **Random hue (add.)** | The contrast (changes within the colour sphere, changing the colour of the image) is change by adding to it a random value within range | 75% | $range = [-0.1, 0.1]$ |
| **Random contrast (mult.)** | The contrast (the brightness range) is change by multiplying it by a random value within range | 75% | $range = [0.7, 1.3]$ |
| **Salt and pepper noise** | Randomly taking pixels and setting them to be 0 (minimum, black) or 1 (maximum, white) | 1% | |
| **Gaussian noise (add.)** | Adding noise to all channels from a normal distribution centered in 0 and with standard deviation | 100% | $\sigma = 0.05$ |

Table S4: Features used for morphometric characterisation.

| Feature (count) | Description | Nuclear (count) |
|---|---|---|
| Area (1) | Area of detected object | X (1) |
| Perimeter (1) | Perimeter of detected object | X (1) |
| Eccentricity (1) | Ratio between major and minor axes | X (1) |
| Circle variance (1) | Measure of the difference between the contour and a circle fitted to the contour | X (1) |
| Ellipse variance (1) | Measure of the difference between the contour and an ellipse fitted to the contour | X (1) |
| Convexity (1) | Ratio between the perimeter of the convex hull of the contour and of the contour | X (1) |
| Solidity (1) | Ratio between the area of the convex hull of the contour and of the contour | X (1) |
| Centroid distance function (CDF) mean, standard deviation, minimum and maximum (4) | Descriptors for the distribution of values in the function characterising the distance between the object center and its edges | |
| CDF noise-insensitive moments (3) | Different ratios between the first four normalised moments ($\mu_1$,$\mu_2$,$\mu_3$,$\mu_4$) of the CDF ($\frac{\sqrt{\mu_2}}{\mu 1}$, $\frac{\mu_3}{\mu_2^{1.5}}$ and $\frac{\mu_4}{\mu_2^2}$) [41] | |
| Curvature function mean, standard deviation, minimum and maximum (4) | Descriptors for the distribution of values in the function characterising the curvature along the contour of the object | |
| Invariant region moments (7) | The first seven region moments. A more detailed description can be found in [41] | |
| CDF Fourier reconstruction error (1) | The area under the curve for the reconstruction error of a Fourier transform of both CDF. In practice: 1) a Fourier transform is fitted to a set of values; 2) the Fourier transform is used to construct a curve of reconstruction errors and 3) the area under the curve for the reconstruction error curve is calculated | |
| Standard deviation, Fourier reconstruction error, minimum and maximum of the intensity profile along the major (4) and minor axis (4) (8 in total) | Descriptors for the distribution of values in the function characterising the intensity profile along the major and minor axis. In practice: 1) the intensity values along an axis are extracted and 2) the standard deviation, Fourier reconstruction error, minimum and maximum are calculated for these values | |
| Mass displacement for the red, green and blue channels and the average intensity (4) | Difference in the center of mass between each colour channel and the average intensity and a uniform prior over the region | X (4) |
| Textural descriptors based on the gray level co-occurrence matrix (GLCM) (4) | Contrast, energy, homogeneity and correlation for GLCM. More details in [41] | |
